# Screening of normal endoscopic large bowel biopsies with artificial intelligence: a retrospective study

**DOI:** 10.1101/2022.10.17.22279804

**Authors:** Simon Graham, Fayyaz Minhas, Mohsin Bilal, Mahmoud Ali, Yee Wah Tsang, Mark Eastwood, Noorul Wahab, Mostafa Jahanifar, Emily Hero, Katherine Dodd, Harvir Sahota, Shaobin Wu, Wenqi Lu, Ayesha Azam, Ksenija Benes, Mohammed Nimir, Katherine Hewitt, Abhir Bhalerao, Andrew Robinson, Hesham Eldaly, Shan E Ahmed Raza, Kishore Gopalakrishnan, David Snead, Nasir M. Rajpoot

## Abstract

**Objectives:** Develop an interpretable AI algorithm to rule out normal large bowel endoscopic biopsies saving pathologist resources.

**Design:** Retrospective study.

**Setting:** One UK NHS site was used for model training and internal validation. External validation conducted on data from two other NHS sites and one site in Portugal.

**Participants:** 6,591 whole-slides images of endoscopic large bowel biopsies from 3,291 patients (54% Female, 46% Male).

**Main outcome measures:** Area under the receiver operating characteristic and precision recall curves (AUC-ROC and AUC-PR), measuring agreement between consensus pathologist diagnosis and AI generated classification of normal versus abnormal biopsies.

**Results:** A graph neural network was developed incorporating pathologist domain knowledge to classify the biopsies as normal or abnormal using clinically driven interpretable features. Model training and internal validation were performed on 5,054 whole slide images of 2,080 patients from a single NHS site resulting in an AUC-ROC of 0.98 (SD=0.004) and AUC-PR of 0.98 (SD=0.003). The predictive performance of the model was consistent in testing over 1,537 whole slide images of 1,211 patients from three independent external datasets with mean AUC-ROC = 0.97 (SD=0.007) and AUC-PR = 0.97 (SD=0.005). Our analysis shows that at a high sensitivity threshold of 99%, the proposed model can, on average, reduce the number of normal slides to be reviewed by a pathologist by 55%. A key advantage of IGUANA is its ability to provide an explainable output highlighting potential abnormalities in a whole slide image as a heatmap overlay in addition to numerical values associating model prediction with various histological features. Example results with can be viewed online at https://iguana.dcs.warwick.ac.uk/.

**Conclusions:** An interpretable AI model was developed to screen abnormal cases for review by pathologists. The model achieved consistently high predictive accuracy on independent cohorts showing its potential in optimising increasingly scarce pathologist resources and for achieving faster time to diagnosis. Explainable predictions of IGUANA can guide pathologists in their diagnostic decision making and help boost their confidence in the algorithm, paving the way for future clinical adoption.

**What is already known on this topic:**

- Increasing screening rates for early detection of colon cancer are placing significant pressure on already understaffed and overloaded histopathology resources worldwide and especially in the United Kingdom^1^.
- Approximately a third of endoscopic colon biopsies are reported as normal and therefore require minimal intervention, yet the biopsy results can take up to 2-3 weeks^2^.
- AI models hold great promise for reducing the burden of diagnostics for cancer screening but require incorporation of pathologist domain knowledge and explainability.

**What this study adds:**

- This study presents the first AI algorithm for rule out of normal from abnormal large bowel endoscopic biopsies with high accuracy across different patient populations.
- For colon biopsies predicted as abnormal, the model can highlight diagnostically important biopsy regions and provide a list of clinically meaningful features of those regions such as glandular architecture, inflammatory cell density and spatial relationships between inflammatory cells, glandular structures and the epithelium.
- The proposed tool can both screen out normal biopsies and act as a decision support tool for abnormal biopsies, therefore offering a significant reduction in the pathologist workload and faster turnaround times.

## Introduction

Histological examination is a vital component in ensuring accurate diagnosis and appropriate treatment of many diseases. In routine practice, it involves visual assessment of key histological and cellular patterns in the tissue, which is a major step in understanding the state of various conditions, such as cancer. Histopathology has been at the forefront of many advances in care including, but not limited to, cancer screening programmes, molecular pathology, tumour classification and companion diagnostic testing, resulting in a rapid rise in demand for histology-derived data^3^. This extra workload is placing tremendous pressure on pathologists, with 78% of UK cellular pathology departments already facing significant staff shortages^4^. The surging demand and staffing challenges ultimately lead to delays in diagnosis, negatively impacting patient care especially for those with abnormal conditions (e.g., cancer or serious inflammation) where early intervention and treatment are critical^5^.

New National Institute for Health and Care Excellence (NICE) guidelines for referral of suspected cancer forecast a rise in demand for endoscopy, with more than 750,000 additional procedures performed per year by 2020^6^, leading to a breach in standard wait times in a quarter of NHS hospitals^7 8^. Endoscopic large bowel biopsies constitute approximately 10% of all requests in the UK National Health Service (NHS) pathology laboratories. During the examination process, the pathologist examines each biopsy slide searching for disease, typically working from low to high magnification, and analyses a set of pre-defined histological features, such as gland architecture, inflammation and nuclear atypia for signs of abnormality^9 10^. The resulting report indicates the presence of any disease process and categorises the abnormality into the most appropriate diagnosis^11 12^. An overview of the pathologist diagnostic decision process for reporting endoscopic colon biopsies is provided in Supplementary Fig 1. Approximately a third of colonic biopsy samples are reported as normal (Supplementary Table 1), representing a substantial workload where the pathologist’s expertise is not fully utilised. The underlying hypothesis of this study is that automated screening of normal biopsies may help address rising histopathology capacity challenges.

Since the advent of digital pathology^13^, there has been a sharp increase in the development of artificial intelligence (AI) tools that enable computational analysis of multi-gigapixel whole-slide images (WSIs). In particular, deep learning (DL) algorithms have achieved remarkable performance not only in routine diagnostic tasks, such as cancer grading^14^ and finding metastasis in lymph nodes, but also in finding origins for cancers of unknown primary (CUP)^15^ and improved patient stratification^16 17^. Notably, Campanella *et al*.^18^ presented a seminal paper on clinical-grade WSI classification, while Bejnordi *et al*.^19^ demonstrated that AI models are capable of surpassing pathologist performance for breast cancer metastasis detection. These models can be leveraged to help reduce inevitable errors in diagnosis, given that humans are naturally prone to mistakes, especially when faced with fatigue or distractions^20 21^. AI tools are not as susceptible to these kinds of errors and therefore may help mitigate oversight, reduce workload and increase reproducibility.

Differentiating between normal and neoplastic colorectal WSIs using DL has previously been addressed, with reports of excellent performance^22-24^. However, distinguishing normal from abnormal tissue samples required for large bowel biopsy screening remains a challenge, due to the difficulty in detecting various subtle conditions, such as mild inflammation. To the best of our knowledge, there are no existing multi-centric studies for normal *vs* abnormal classification of large bowel biopsies. Existing methods for colonic analysis operate on high power sub-images (or image patches) and so do not explicitly model both the tissue micro- and macro-structure, including glandular architecture, inflammatory cell density and spatial relationships between inflammatory cells, glandular structures and the epithelium. Relying solely on DL models to automatically detect histological patterns that are diagnostically relevant in small image regions may lead to sub-optimal performance. Alternatively, explicitly incorporating histological features that are routinely used by pathologists during the colon biopsy diagnostic workflow may not only improve performance over conventional DL models but may also increase transparency and interpretability of the algorithm’s decision making to the pathologist – a key requirement for trustworthy AI based medical decision models^25 26^.

To help reduce the burden of large bowel biopsy screening, we propose the first interpretable AI algorithm for large bowel slide classification employing a gland-graph network named IGUANA (Interpretable Gland-Graphs using a Neural Aggregator). In the proposed approach, a WSI is modelled as a graph with nodes^27-30^, each representing a gland associated with a set of 25 interpretable features capturing gland architecture, intra-gland nuclear morphology and inter-gland cell density. The interconnections between these nodes capture the spatial organisation of glands within the tissue. The node features were developed in collaboration with pathologists and in accordance with existing diagnostic pathways to boost predictive accuracy, interpretability and alignment with known histological characteristics of a wide range of colorectal pathologies. IGUANA identifies highly predictive regions in the biopsy tissue slide and provides an explanation as to *why* they may be highly predictive. Because of the use of biologically meaningful features, this explanation can easily be interpreted by a pathologist as the basis of the algorithm’s diagnostic decision making. We validate our algorithm on an internal dataset containing 5,054 WSIs and an independent multi-centre dataset containing 1,561 WSIs, achieving the best performance compared to recent top-performing approaches. In addition, we analyse predictive regions identified by IGUANA along with local and WSI-level explanations and show that our approach can identify areas of abnormality, such as inflammation and neoplasia. A summary of our overall pipeline can be seen in Fig 1, which consists of the following steps: 1) histological segmentation, 2) feature extraction and edge generation, 3) graph prediction and 4) graph explanation. The code for IGUANA is available in the open-source domain for research purposes (https://github.com/TissueImageAnalytics/iguana) and example results can be visualised in an interactive demo available at https://iguana.dcs.warwick.ac.uk.

**Fig 1:**
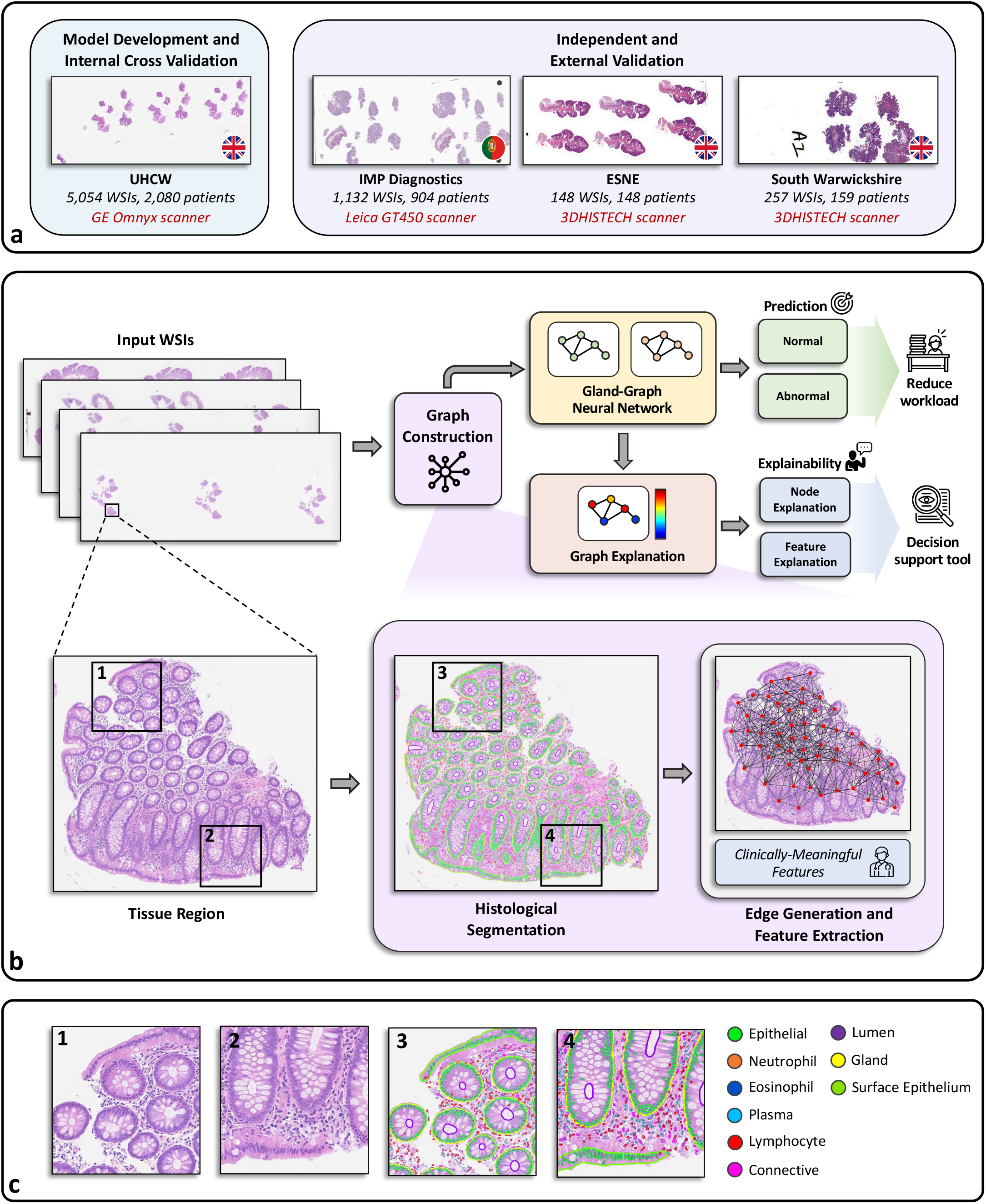
Illustration of the overall pipeline for colon tissue classification with gland-graph convolutional networks. **a**, Overview of the data used in our experiments from 4 different centres utilizing different scanners. **b**, Summary of the pipeline, which involves graph construction, gland-graph inference and gland-graph explanation. **c**, Zoomed-in image regions and corresponding results taken from the example in panel **b**.

## Methods

### WSI datasets

We collected data from four patient cohorts containing routine Haematoxylin and Eosin (H&E) stained WSIs of endoscopic colon biopsies from the following centres: 1) University Hospitals Coventry and Warwickshire (UHCW) NHS Trust, United Kingdom; 2) South Warwickshire NHS Foundation Trust, United Kingdom; 3) East Suffolk and North Essex (ESNE) NHS Foundation Trust, United Kingdom and 4) IMP Diagnostics Laboratory, Portugal^23^. Glass slides from UHCW were digitised with a GE Omnyx slide scanner at a pixel resolution of 0.275 microns per pixel (MPP). Slides from ESNE and South Warwickshire Hospitals were digitised with 3DHISTECH scanners at pixel resolutions of 0.122 MPP and 0.139 MPP, respectively. IMP Diagnostics slides were digitised with a Leica GT450 scanner at a pixel resolution of 0.263 MPP. In total, we collected 6,591 WSIs from 3,291 patients, with 5,054 from UHCW, 148 from ESNE, 257 from South Warwickshire and 1,132 from IMP Diagnostics. To rigorously evaluate our approach for colon biopsy screening, we performed 3-fold internal cross-validation on the UHCW dataset and held out the remaining three datasets for independent external validation. When creating the folds for internal cross-validation, the data was split with stratification at patient-level to ensure that our method was evaluated on completely unseen cases.

Collaborating pathologists categorised WSIs from UHCW, ESNE and South Warwickshire at slide-level into a ground-truth diagnosis label of either normal, non-neoplastic or neoplastic with consensus review of discordant cases. A wide range of histological conditions were present across the datasets to reflect the clinical screening procedure. A full summary of the specific diagnoses is shown in Supplementary Table 2. For this study, non-neoplastic and neoplastic classes were combined into a single *abnormal* category. WSIs from IMP diagnostics were originally categorised as either non-neoplastic, low-grade dysplasia or high-grade dysplasia^23^, where the non-neoplastic category contained a mixture of normal, inflammatory and hyperplastic slides. Therefore, our team of pathologists additionally reviewed non-neoplastic slides from IMP to separate normal from abnormal tissue samples. In our final curated datasets, 42% of slides from UHCW, 61% from ESNE, 40% from South Warwickshire and 84% from IMP Diagnostics were labelled as abnormal. We provide a data description diagram showing the experiment design and the inclusion and exclusion criteria used in Supplementary Fig 2. In addition, we provide an overview of all datasets used in this study in Supplementary Fig 3 and give a demographic breakdown of patients within the development set in Supplementary Fig 4.

### Identification of histological objects

The first step of IGUANA requires the segmentation of various histological objects within the WSI, which enables subsequent graph construction and feature extraction. For this, we utilise our recently published *Cerberus*^31^ model, which performs simultaneous segmentation and classification of nuclei, glands, lumen and different tissue regions. During training, we use a multi-task learning strategy, which allows the utilisation of multiple independent datasets and enables simultaneous prediction with a single network. Therefore, our localisation step is computationally efficient and does not require multiple passes through various networks. As well as delineating object boundaries, Cerberus determines the category of each nucleus and differentiates the surface epithelium from other glands. Cerberus is trained on a large amount of data from 12 different centres, including more than 535K nuclei, 51K glands and 56K lumen annotations. In our previous work, we have shown that this crucial step of initial localisation generalises well to unseen examples^31^.

### Extraction of clinically interpretable features

After performing segmentation of various histological objects using *Cerberus*, we extract a set of clinically meaningful features, which were carefully chosen in collaboration with pathologists so that they reflect what features are considered during the screening procedure. Our model’s ability to localise glands, lumen and nuclei within the tissue allows us to extract interesting gland, intra-gland and inter-gland features that are potentially capable of identifying various histological conditions. Specifically, the inter-gland features are defined in the non-glandular surrounding area, also known as the *lamina propria*. To obtain this region, we utilise the patch-based tissue type classification output from Cerberus and consider both normal gland and tumour patch predictions. We then subtract the gland segmentation output from the prediction map and carry out a series of refinement steps to obtain the final estimation of the lamina propria. We ensure that each feature that we consider has a key biological significance. For example, we consider the size and morphology of glands, which can be indicative of cancer. For quantifying the morphology, we utilise the best alignment metric (BAM)^32^, which provides a measure of how elliptical an object is, to help capture abnormal glands with irregular shapes. We also take into account the number of lumen along with their corresponding morphology, which can be suggestive of conditions such as cribriform architecture and serrated polyps, respectively. Furthermore, the organisation of epithelial cells and the amount of different inflammatory cells within the gland are diagnostically informative. For example, normal glands will have epithelial cells organised at the boundary and neutrophils within the gland are indicative of crypt abscesses. For measuring the epithelial organisation, we compute the mean and standard deviation of distances of epithelial nuclei centroids to their nearest gland boundary. We also utilise the mean and standard deviation of inter-epithelial nuclear distances within the gland. Certain inflammatory conditions, such as lymphocytic colitis, will have an increased number of inflammatory cells within the lamina propria. Therefore, we extract inter-glandular features indicative of the local density of inflammatory cells and report the associated cellular composition. Overall, we compute a set of 25 features, which are standardised before utilisation within our graph-based machine learning model. Visual examples of features used within our framework, along with examples from the 5^th^ and 95^th^ percentiles, are given in Supplementary Fig 5. We also provide a more in-depth description of these features, along with what conditions they can detect in Supplementary Table 3.

### Gland-graph neural network for interpretable diagnosis

Recently, graph neural networks (GNNs) have become popular in Computational Pathology (CPath)^27^ due to their ability to model a large WSI as an interconnection of nodes representing histologically important constructs characterized by node-level features^28 33 34^. An added advantage of using GNNs for predictive modelling in CPath is their ability to generate an explanation of their output in terms of the node level features^35-37^.

Thus, once the different histological objects have been identified, each WSI is represented as a gland-graph. Here, glands are represented as nodes on a 2D plane that are connected by edges if they are within a certain distance of each other. Each gland is then associated with a set of 25 features that were previously described. Therefore, the overall graph provides a mechanism for representing local features across the entire tissue sample. As opposed to surgical resections, which usually contains a large bulk of tissue, biopsies can contain many separate tissue segments on the slide. This arrangement has no biological significance and, therefore, it would be unreasonable for glands to be connected between neighbouring tissue regions. Thus, we also ensure that an edge between any two given glands only exists if they are both located within the same tissue segment.

Upon formation of our gland-graph representation of a WSI in terms of its nodes and edges, we pass the input through a GNN, which sequentially aggregates features within the slide to predict the diagnosis. An important aspect of IGUANA is its ability to provide an interpretable and explainable output, which can be used to facilitate the diagnostic process and potentially for biomarker discovery. For this, we utilise GNNExplainer^36^, which generates a subset of nodes and features that play a crucial role in the GNN’s prediction. To obtain a WSI-level explanation, the local features are averaged within the top ten most predictive nodes. This enables analysis over larger cohorts to identify existing sub-populations. To further increase model interpretability, we can also visualise the intermediate nuclear, lumen and gland localisation results overlaid on top of the original WSI. We provide further detail of our graph-based approach in Supplementary Section S3.

### Software, optimisation and reproducibility

We implemented our framework with the open-source software library PyTorch version 1.10^38^, PyTorch Geometric version 2.1.1^39^ and Python version 3.6 on a workstation equipped with one NVIDIA Tesla V100 GPU. We utilised scikit-learn version 1.0.2^40^ to perform the comparative experiments using random forest and fastcluster version 1.2.6^41^ to perform biclustering analysis. We trained our graph neural network for 50 epochs using a batch size of 64 and an initial learning rate of 0.005, which was reduced by a factor of 0.2 after 25 epochs. It should be noted, that despite using a GPU with 32GB RAM, our GNN framework incurred a low memory utilisation and therefore different specification GPUs may also be used. The interactive demo was developed using the tile server from TIAToolbox^42^ and Bokeh 2.4.3. No changes were made to the AI system or the hardware over the course of the study.

Model code, along with a full list of software requirements, is located at https://github.com/TissueImageAnalytics/iguana. Model code and weights are for research purposes only and are therefore shared under a non-commercial Creative Commons license.

### Patient and public involvement

Lay members have made a valuable contribution to this project in ensuring that the patient is at the heart of this project. Three lay advisors have been working with us since the conception of this project. One of the advisors is part of the National Cancer Research Institute (NCRI) consumer network and Independent Cancer Patient’s Voice (ICPV) group, who are both supportive of new technologies being brought into the NHS for patient benefit.

## Results

### Large-scale cross validation for colon biopsy screening

To rigorously evaluate our approach for colon biopsy screening, we performed 3-fold cross-validation using 5,054 Haematoxylin and Eosin (H&E) stained colon biopsy WSIs from University Hospitals Coventry and Warwickshire (UHCW), where each slide was labelled as either normal or abnormal. Interpretable screening of normal colon biopsies is a challenging problem due to a wide spectrum of large bowel abnormalities including a variety of neoplastic and inflammatory conditions. Fig 3 shows the results of IGUANA, achieving an average area under the receiver operating characteristic (AUC-ROC) curve of 0.9783 ± 0.0036 and an area under the precision-recall (AUC-PR) curve of 0.9798 ± 0.0031. These scores determine the level of agreement between consensus pathologist diagnosis and AI generated classification of normal versus abnormal biopsies. We also include results obtained using other existing slide-level classification algorithms such as IDaRS^43^, CLAM^44^ and a random forest (RF) baseline classifier using our glandular features (denoted by Gland-RF). We observe that IGUANA achieves the best performance compared to both patch-based methods (IDARS and CLAM), demonstrating its strong predictive ability given that it uses only 25 features per gland. We provide additional comparative results between IGUANA and IDaRS in Supplementary Fig 6. Note that despite IGUANA outperforming it, the Gland-RF model produces comparable performance – signifying the strength of our set of clinically-derived features – albeit without the localised interpretability provided by IGUANA. Also, as opposed to the two patch-based methods, IGUANA provides concrete justification as to *why* a certain diagnostic class was predicted. We go into further detail on interpretability and explainability later in this section.

### Model generalization to independent cohorts

A true reflection of a model’s clinical utility requires the assessment of its performance on completely unseen cohorts. For this, we utilised three additional cohorts of H&E-stained colon biopsy slides, providing a total of 1,537 WSIs. These cohorts consisted of 1,132 slides from IMP Diagnostics Laboratory in Portugal^23^, 148 slides from East Suffolk and North Essex (ESNE) NHS Foundation Trust and 257 slides and South Warwickshire NHS Foundation Trust, where slides were again categorised as either normal or abnormal. We observe from Fig 3 that our model attains high performance for both the ESNE and South Warwickshire cohorts, reaching AUC-ROC scores of 0.9567 ± 0.0155, 0.9649 ± 0.0025 and 0.9789 ± 0.0023 and AUC-PR scores of 0.9731 ± 0.0105, 0.9466 ± 0.0034 and 0.9949 ± 0.0006 for ESNE, South Warwickshire and IMP datasets, respectively. It is evident that there is a large difference in performance between IGUANA and other approaches on the external cohorts, signifying that superior generalisation to unseen data is a strength of our model. In particular, at a sensitivity of 0.99 we obtain a percentage increase over IDaRS of 47.4%, 63.6% and 58.9% for IMP, ESNE and South Warwickshire cohorts, respectively. Example results obtained by our segmentation model across the four datasets are shown in Fig 2.

**Fig 2:**
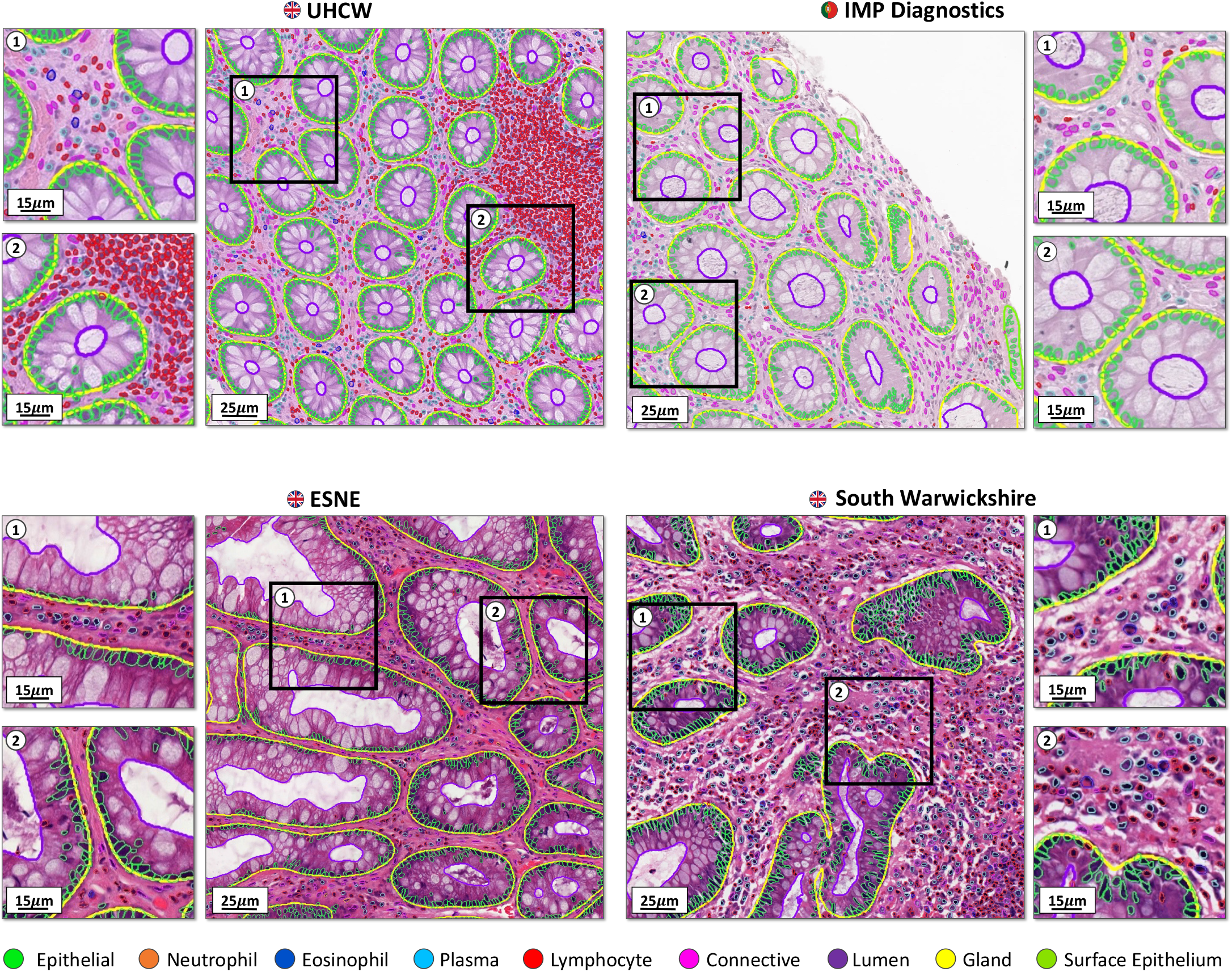
Example segmentation results obtained by our multi-task model across the four datasets used in our experiments. The top row shows normal examples, whereas the bottom row shows abnormal examples. In particular, the bottom-left example from ESNE shows a hyperplastic polyp and the bottom-right example from South Warwickshire shows inflammation.

**Fig 3:**
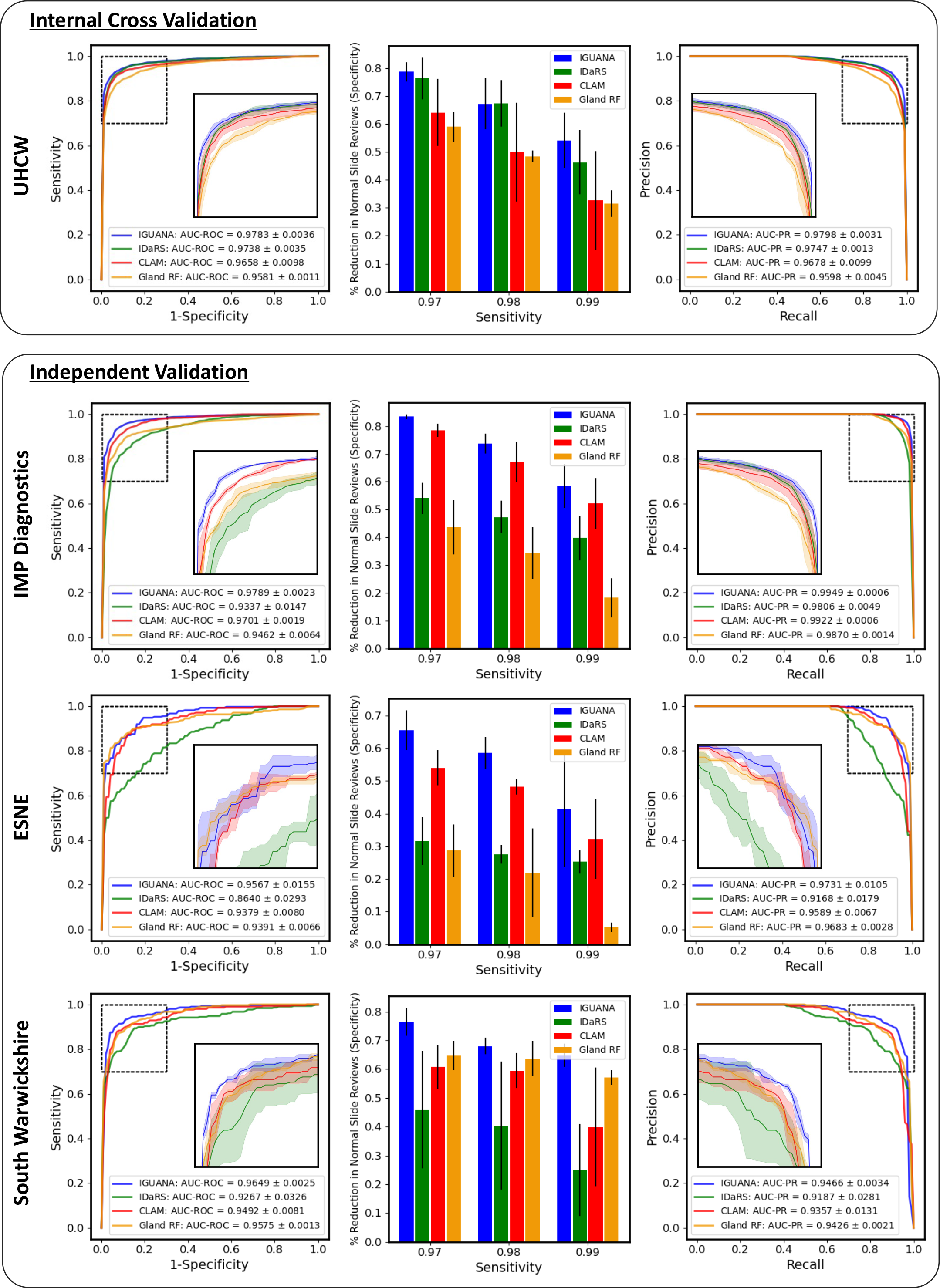
Results obtained across the 4 cohorts used in our experiments. Here, we display the ROC and PR curves along with the respective AUC scores of our approach compared to IDaRS, CLAM and Gland-RF (a random forest approach using the same handcrafted features with global aggregation). We also display the specificities obtained at sensitivity cut-offs of 0.97, 0.98 and 0.99. The shaded areas in the curves and the error bars in the bar plots show 1 standard deviation from the results.

### Analysis of expected reduction in pathologist workload

The real-world value of our approach is determined by its ability to reduce pathologist workload. As our model is intended for screening, it must achieve high sensitivity to minimise the risk of false negatives. Therefore, assessment of the specificity at high sensitivity cut-off thresholds provides a good indication of its potential effectiveness as a screening tool. Here, the specificity is directly indicative of the percentage reduction in normal slides that require pathologist review. In the middle column of Fig 3 we display the specificity of our model at sensitivities of 0.97, 0.98 and 0.99 on all datasets used in our experiments, where we see that IGUANA sustains the best performance at various cut-offs compared to other methods. During internal cross-validation, we obtain specificities of 0.7865 ± 0.0429, 0.6720 ± 0.1128 and 0.5409 ± 0.1210 for sensitivities of 0.97, 0.98 and 0.99, respectively. For independent validation, our method obtains average specificities across the three external datasets of 0.7513 ± 0.0919, 0.6679 ± 0.0779 and 0.5487 ± 0.1599 for sensitivities of 0.97, 0.98 and 0.99. Therefore, this indicates that at a sensitivity of 0.99, our method is able to screen around 55% of normal cases during both internal and external validation.

In Supplementary Fig 7 we show the proportion of slides that require pathologist review to achieve a certain sensitivity^18^. In these plots, we consider a target sensitivity of 0.99, which is reasonable due to high levels of inter-observer disagreement for conditions such as mild inflammation. We also show with a vertical dashed line the proportion of abnormal slides in each dataset, which indicates the minimum number of slides that need to be reviewed for screening. For each of the cohorts, we observe that for our target of 0.99 sensitivity our model can screen out 32%, 31%, 17% and 13% of total slides from UHCW, South Warwickshire, ESNE and IMP datasets, respectively. If considering a sensitivity of 0.97, we can screen out 44% of slides from UHCW, 46% from South Warwickshire, 30% from ESNE and 19% from IMP.

### Performance across anatomical sites

We perform a statistical analysis of test results on the internal UHCW dataset to analyse potential differences in performance over colonic sites, such as the ascending, descending and transverse colon. After performing the Mann-Whitney U test^45^ between the results of different sites, we found that no statistically significant difference was observed, with *p*-values > 0.05. This suggests that there is insufficient evidence of significant differences in the results across anatomical sites.

### Local feature explanations increase model transparency

A major component of IGUANA is the ability to provide an interpretable and explainable output. In Fig 4, we display visual explanations of the most predictive nodes and features given by IGUANA. Node explanations are shown in the form of a heatmap, where relatively high values indicate glandular areas that contribute to the slide being classified as abnormal. Therefore, we should expect that all glands in a normal slide will have low values in the associated heatmap as shown in Fig 4*a*, where no glands contribute to the slide being classified as abnormal. Fig 4*b-d* show WSIs with hyperplastic polyps, inflammation and adenocarcinoma, respectively. Hyperplastic polyps are often characterised by intraluminal folds and lumen dilation. On the other hand, inflammatory conditions usually have an increased number of lymphocytes, plasma cells, eosinophils and neutrophils within the lamina propria and potentially within the glands. Other indicators of inflammation can include crypt branching and crypt dropout. Colon adenocarcinoma is often denoted by irregular glandular morphology, epithelial nuclear atypia and multiple lumina. High-grade cancers typically lose their glandular appearance and form solid sheets of tumour cells. It can be observed that IGUANA is able to pick up abnormal glands with features in line with the above descriptions. In particular, we see that the most predictive glands in Fig 4*b* contain lumen with a clearly irregular morphology, whereas highlighted glands in 4*c* show areas with a high degree of inflammation. The adenocarcinoma heatmap in Fig 4*d* highlights areas that have lost their conventional glandular appearance. Specifically, epithelial nuclei are no longer arranged at the gland boundary, cribriform architecture is observed and glands appear much larger, due to the formation of tumour cell sheets.

**Fig 4:**
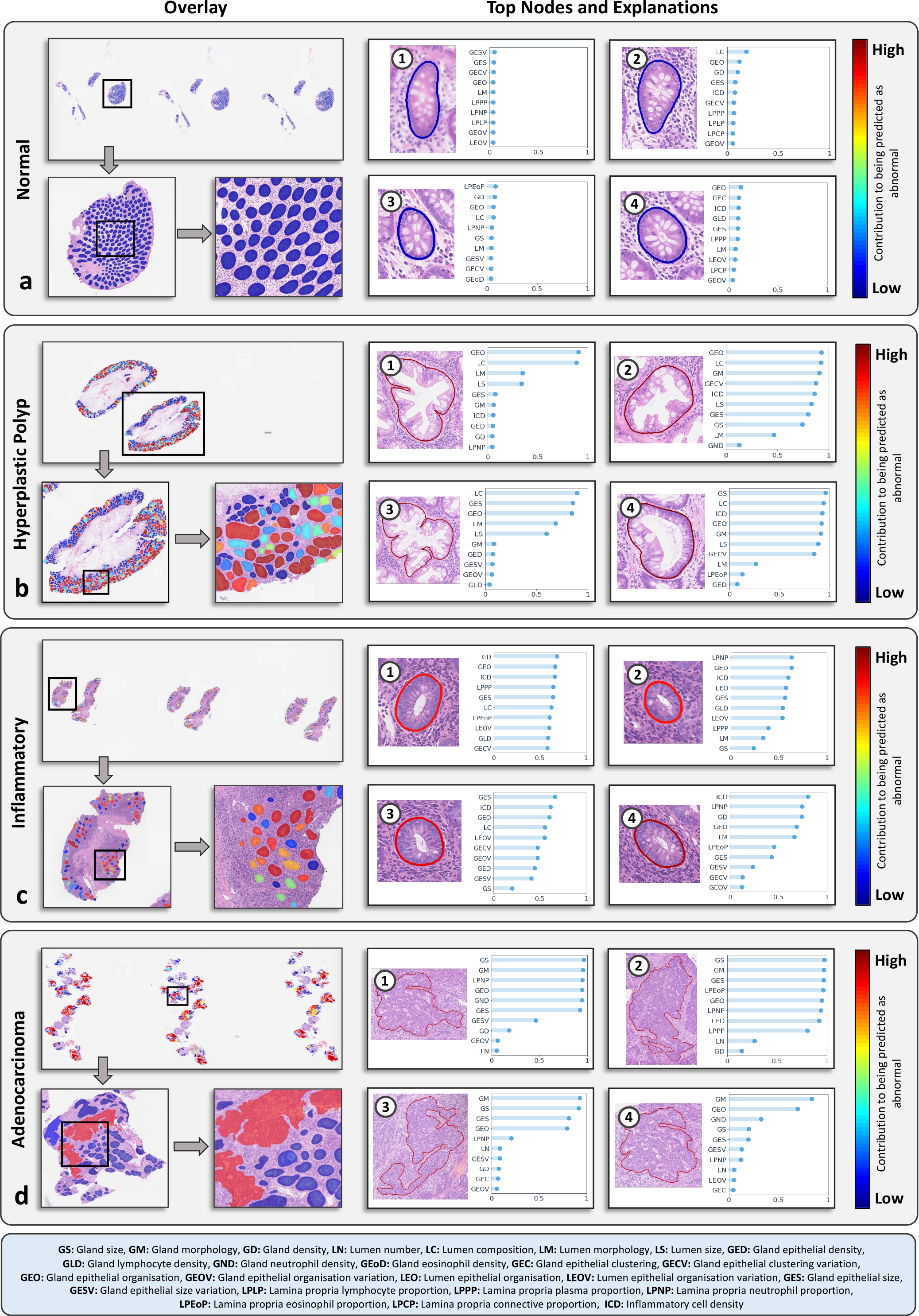
Visualisation of node and feature explainability. Here, we present the overlay of the node-level explanations in the form of a heat map which show the most predictive nodes in the WSI. We also show cropped images of the 4 most predictive nodes within each WSI along with the associated 10 most predictive features and their feature importance value. The colour of the boundary of the top nodes (glands) indicates the corresponding value in the node explanation heatmap. **a**-**d** show example slides that are normal, hyperplastic, inflammatory or cancerous, respectively.

In addition to the node explanation heatmap, IGUANA indicates *why* certain glands are being identified as abnormal. This is useful because it can provide confirmation that the correct features are being identified by the model, giving researchers and clinicians confidence that it is performing as expected. This strategy can also be used to identify additional features within abnormal conditions. To show this, in Fig 4, we display the most predictive glands in each slide and provide the corresponding feature explanations. Specifically, we display the top ten features in descending order of significance, along with their corresponding feature importance values between 0 and 1. Here, we expect that the feature explanations should align with what is observed in the associated cropped regions. In our hyperplastic polyp example, we see that the top glands (i.e., 1, 2 and 3) contain lumen with abnormal morphology, whereas lumen dilation is observed in top gland 4. In line with this, lumen morphology and lumen composition are high-scoring features across the provided examples. We also observe that lumen size and organisation of epithelial nuclei within the glands are often found to be important features. In the example shown in Fig 4*c*, we observe that top glands have a high degree of inflammation, which is matched by top features, such as inflammatory cell density, gland density and lamina propria neutrophil proportion. In the adenocarcinoma example, we see that the top four glands are all large, have irregular morphology and often display solid sheets of tumour cells with no obvious glandular structure. This is highlighted in the feature explanation, where gland morphology, gland size and epithelial organisation are consistently top-ranked features. Here, epithelial organisation describes how the epithelial nuclei are positioned at the gland boundary. Due to the presence of solid tumour patterns across the top glands, this feature is frequently highlighted in cancerous cases. We provide additional visual examples of the interpretability of our model output in Fig 5.

**Fig 5:**
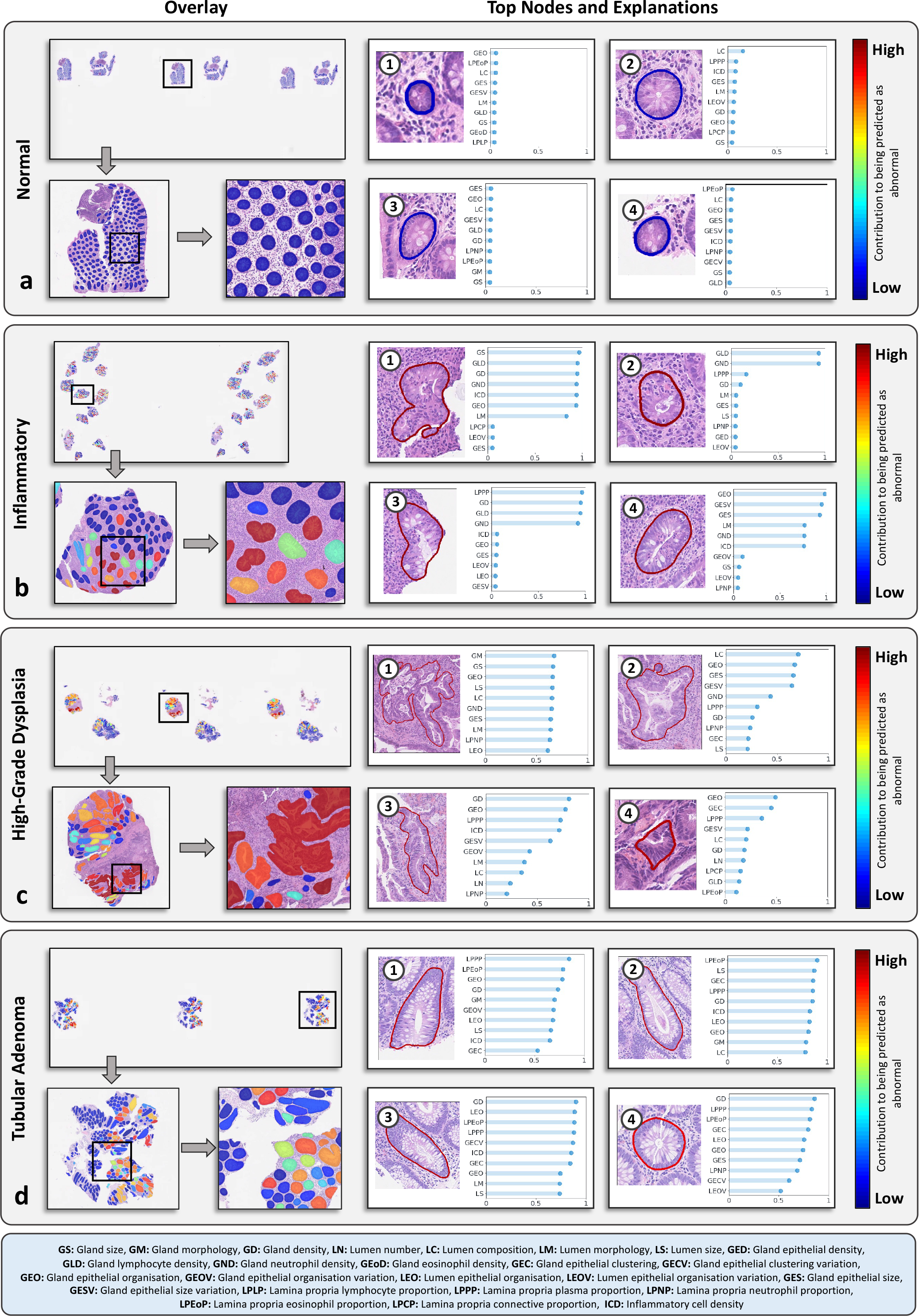
Additional visualisation of node and feature explainability. As before, we present the overlay of the node-level explanations in the form of a heat map which show the most predictive nodes in the WSI. We also show cropped images of the four most predictive nodes within each WSI along with the associated 10 most predictive features and their feature importance value. **a**-**d** contain show slides that are normal, inflammatory (with crypt abscesses), high-grade dysplasia or adenomatous polyps, respectively.

### WSI-level feature explanations are consistent with known histological patterns

In Fig 6*a*, we show WSI-level explanations averaged over different sub-conditions in the UHCW and IMP cohorts. We focus on these datasets because they are the largest, with both containing over 1,000 samples. Here, we plot top 10 features across the various sub-conditions for increased readability. These plots can be used both to confirm that the global explanations are as expected and to understand which features are particularly important for categorising a certain sub-condition as abnormal. In both UHCW and IMP cohorts, the normal radar plots have a small radius, indicating that no feature contributes to the slide being classified as abnormal. For inflammatory cases, the UHCW and IMP radar plots show that a wide range of features can contribute to the slide being classified as abnormal, where there may be both cellular and architectural changes in the tissue. However, the most important features that can differentiate between other sub-conditions include inflammatory cell density, gland lymphocyte infiltration and gland density. Gland density can be indicative of gland dropout, which is a sign of inflammation. The UHCW radar plots for dysplasia and adenocarcinoma are similar, where the most important features are gland morphology, gland epithelial cell organization, gland epithelial cell size and variation of gland epithelial cell size. This is in line with the key expected histological patterns observed within these tissue types. Likewise, these plots are similar to the low- and high-grade dysplasia plots for the IMP cohorts, indicating that the correct histological features are being highlighted when providing the WSI feature explanation. For hyperplastic polyps, we can see that lumen composition, lumen morphology and epithelial cell organisation have a large influence in the slide being classified as abnormal. Lumen composition is the ratio of lumen to gland size and therefore can identify glands with lumen dilation, which is a distinguishing feature of hyperplastic polyps. Conversely, lumen serrations, which are present in hyperplastic polyps, can lead to irregular lumen morphology, further validating the feature explanations output by our model.

**Fig 6:**
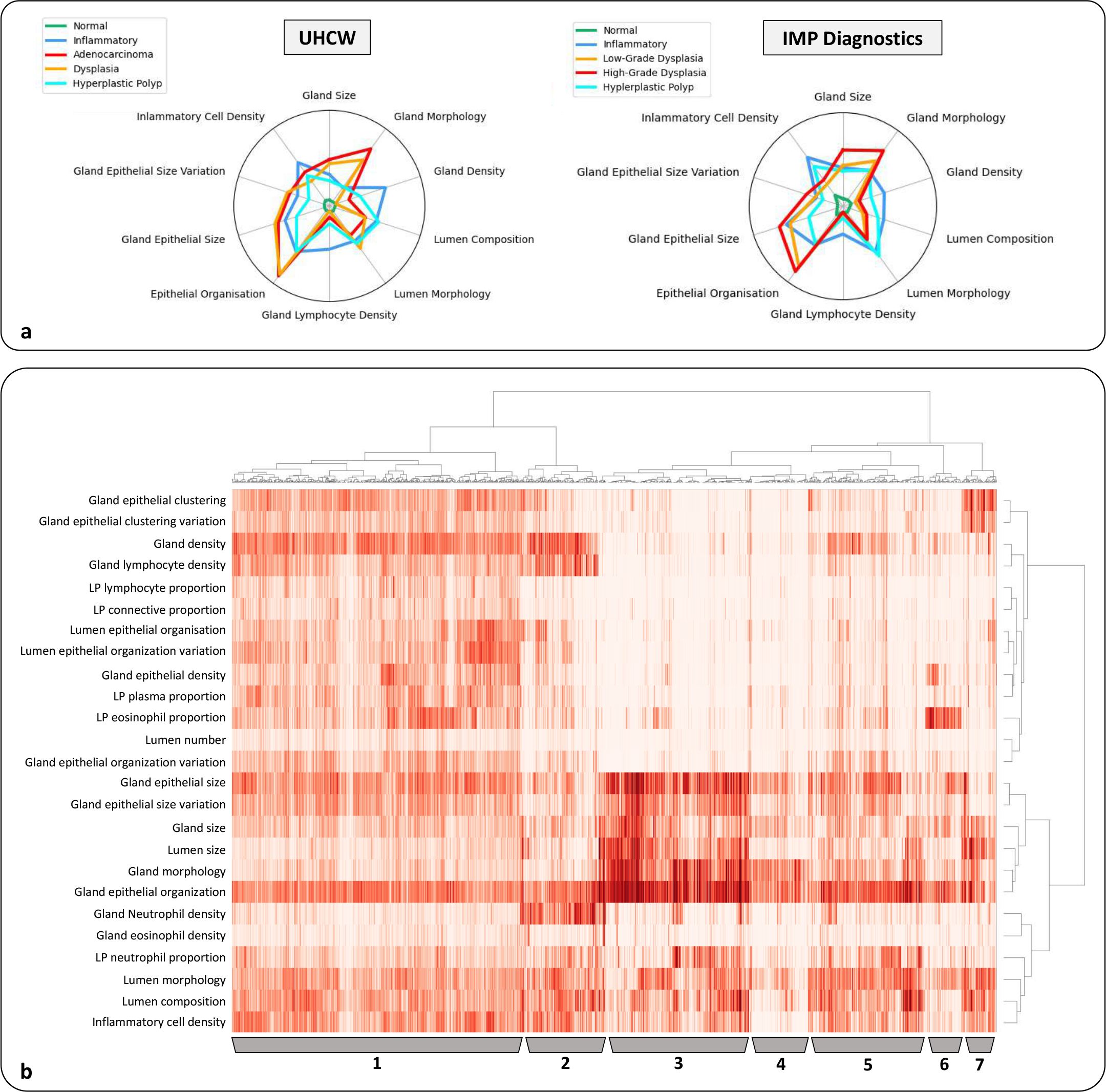
Analysis of global explanations. **a**, Radar plots showing global feature importance for sub-conditions in the UHCW and IMP datasets. **b**, Hierarchical biclustering of feature importance values. 1-7 denote prominent clusters after biclustering, with the following distinguishing histological characteristics: 1) inflammation, without gland neutrophil infiltration; 2) inflammation with both gland lymphocytic and neutrophilic infiltration; 3) neoplasia with irregular gland morphology and large epithelial cells; 4) irregular gland morphology with minimal inflammation; 5) hyperplasia with irregular lumen morphology and composition with inflammation in the lamina propria; 6) eosinophilic infiltration in the lamina propria and 7) neoplasia with gland epithelial cell clustering.

### WSI-level feature explanations identify population subgroups

In Fig 6*b*, we perform hierarchical biclustering of all abnormal slides and WSI-level feature importance scores to help identify various subgroups that exist within the UHCW dataset. At the bottom of the plot, we identify various patient clusters which have varying histological appearance. These are numbered as follows: 1) general sign of inflammation, without neutrophil infiltration; 2) inflammation with a high degree of both lymphocytic and neutrophilic gland infiltration; 3) mainly neoplastic slides with irregular-shaped glands and large epithelial cells; 4) irregular gland morphology, with minimal inflammation; 5) abnormal lumen morphology and composition, with signs of inflammation in the lamina propria; 6) increased eosinophilic infiltration in the lamina propria and 7) neoplastic slides with gland epithelial clustering. Therefore, this gives us confidence that the network is learning key histological differences among the dataset to make an informed WSI-level prediction. More fine-grained clusters can be observed by referring to the associated dendrograms in the biclustering plot.

### Interactive visualisation of results

We provide an interactive demo at https://iguana.dcs.warwick.ac.uk showing sample IGUANA results and highlighting the full output of our model at global and local levels, including the intermediate gland, lumen and nuclear segmentation results. In particular, we display the node explanations overlaid as a heat map on top of the glands and the local explanations by hovering over each node in the overlaid graph. Here, we provide the top 5 features to provide insight into what is contributing to certain glands being flagged as abnormal. It may also be of interest to assess the difference in features for nodes across the WSI. Therefore, we also enable visualisation of each of the 25 features overlaid on top of the glands as heat maps.

## Discussion

### Key findings

There has been a staffing crisis in pathology for many years^46^, which is being further exacerbated by the increased demand for histopathological examination. Embracing new technologies and AI in clinical practice may be necessary as hospitals seek to find new ways to improve patient care^47^. AI screening of large bowel endoscopic biopsies holds great promise in helping to reduce these escalating workloads by filtering out normal specimens. However, currently there doesn’t exist a solution that can do this with a high predictive performance. Also, explainable AI is now recognised as a key requirement for trustworthy AI in human-centred decision making^26^, yet is usually not considered in many healthcare applications. Therefore, in this study we developed an AI model that can accurately differentiate normal from abnormal large bowel endoscopic biopsies, while providing an explanation for why a particular diagnosis was made.

We demonstrated that our proposed method for automatic colon biopsy screening could achieve a strong performance during both internal cross validation (mean AUC-ROC=0.98, mean AUC-PR= 0.98) and on three independent external datasets (mean AUC-ROC=0.97, mean AUC-PR=0.97). Highly sensitive tools for screening are required to minimise the number of undetected abnormal conditions, since the false negative report is likely to lead to delayed diagnosis and potential patient harm. Currently, we obtain promising specificities of 0.789 ± 0.043 at a sensitivity of 0.97 and 0.541 ± 0.121 at a sensitivity of 0.99, which could have a positive impact on reducing pathologist workloads. We also show in Supplementary Fig 7 the expected reduction in clinical workload, where we report up to a 32% time saving by screening out normal biopsies that do not require assessment, while still maintaining a sensitivity of 0.99.

### Analysis of misclassifications

To understand misclassifications made by our model, we show six normal slides with the highest predicted abnormality scores in Supplementary Fig 8. After inspection, we see that IGUANA correctly classifies these slides and therefore identifies mislabelling errors in the dataset. Here, the examples should have been labelled as either inflammatory or hyperplastic polyp. In the figure, we include sample image regions, as well as local and WSI-level feature explanations that are reflective of the true category of each slide. In addition, we performed a false negative analysis, where in Supplementary Fig 9*a* we show the counts of various sub-conditions along with the corresponding number of false negatives. In Supplementary Fig 9*b*, we show the false negative rate of each category. It can be observed that the model found slides with lymphocytic and collagenous colitis somewhat challenging, with false positive rates of 0.29 and 0.46, respectively. Explicit modelling of the sub-epithelial collagen band should enable us to better detect collagenous colitis. It may be worth noting that there was a relatively small number of collagenous colitis samples in all four cohorts and so they may not have a large impact on the overall performance. Also, a high false negative rate was observed in the mild inflammation category, but this is to be expected because they are visually similar to normal samples.

### Strengths and limitations

Not only does the IGUANA algorithm attain a strong performance, it also highlights diagnostically important areas within the tissue and provides an explanation as to which clinical features in those areas are most related to its prediction. This is a major strength of our study as it implies that IGUANA can be used to screen out normal biopsies with high accuracy and also used as a highly interpretable decision support tool within abnormal biopsies.

To enable explainable predictions, our algorithm relies on an accurate intermediate segmentation step, which requires many pixel-level annotations. This can be a time-consuming step and can therefore act as a bottleneck in the development of similar methods. In addition, the type of features that can be incorporated into our AI algorithm are dependent on which kinds of histological objects are initially localised. For example, we do not currently detect goblet cells and so do not include features indicative of goblet cell-rich hyperplastic polyps. Other histological objects that could be added include giant cells, signet ring cells and mitotic figures. In addition, although we segment the surface epithelium, we do not extract any associated features that can help identify conditions such as collagenous colitis. Our method also does not assess surface abnormality to detect intestinal spirochaetosis or pigment to detect melanosis coli. These shortcomings will be addressed in future work. In Supplementary Fig 1*b*, we highlight diagnostic features (in red colour) that are not currently modelled in our framework.

### Findings in context

There have been recent AI approaches developed for cancer detection in colonic whole-slide images^22 48 49^. However, such approaches cannot be used for screening in clinical practice because they often fail to identify non-cancerous abnormalities such as inflammation. Similarly, AI models have been developed for detecting polyps^50 51^, inflammatory bowel disease^52^ or grading dysplasia^23^, but again they do not address the problem of screening normal from all types of abnormality. Our approach utilises retrospective biopsies from pathology archives, where data is accordingly labelled as normal or abnormal to reflect the clinical screening process. Therefore, unlike other approaches, our AI model can be directly implemented as a triaging tool and may therefore have a profound effect on reducing pathologist workloads. In addition, most recent automatic methods rely on weak supervision, where only the overall diagnosis is used to guide the algorithm. This strategy may be advantageous because it does not rely on the time-consuming task of collecting many annotations. However, this limits the interpretability of the output, which may hinder the acceptance of such models in hospitals.

### Implications for clinicians and policy makers

Analysis of colon biopsy slides by visual examination, either under the microscope or more recently on the computer screen, is the current gold standard. However, the current practice is unsustainable with increasing numbers of specimens that require examination and due to staff shortages, where only 3% of NHS hospitals report adequate staffing^4^. With advances in cancer screening programmes and no immediate sign of the pathologist staffing crisis being resolved, additional measures to assist with reporting will be essential. Our proposed AI model addresses this unmet need by automatically filtering out normal colonic biopsies that require minimal intervention, yet make up a substantial proportion of all cases, with high degree of accuracy. As a result, our model significantly reduces the number of samples that require review by pathologists.

AI models are now starting to be used in clinical practice for prostate cancer detection, where a clear advantage for clinicians has been demonstrated in terms of reducing workloads and increasing reporting accuracy^53 54^. There is growing evidence that automated methods for tissue diagnosis can transform pathologist workflows and help drive new policies in healthcare. However, no such tool currently exists for screening large bowel endoscopic biopsies, perhaps due to the fact that no automated tool has been able to accurately detect all kinds of abnormality, including inflammation, dysplasia, hyperplasia and neoplasia. With its triaging capability, the proposed model promises to have positive implications on patient treatment due to faster time to diagnosis, resulting in the potential for early intervention where it is needed the most.

### Extending the model to surgical resections

Despite the screening of endoscopic large bowel biopsies being a focus of this study, the proposed approach could be applied to resection samples with minimal modification. Our method may be especially powerful in this case because each tissue segment within the slide is typically larger, allowing greater spatial context to be explored. Two potential areas of interest using resection samples include the prediction of genetic alterations^55^ and survival analysis^17 56^. In these cases, histological biomarkers are less well known, as compared to those used for routine screening tasks. Therefore, our approach might be used to aid biomarker discovery and help toward further understanding of which morphological patterns are associated with certain genetic alterations and clinical outcomes.

## Conclusion

We have shown that IGUANA offers promise as an effective triaging tool for AI-based colon biopsy screening with a strong emphasis on diagnostic interpretability providing concrete justification as to *why* a certain diagnostic class was predicted and making its predictions transparent and explainable. The proposed AI method can help alleviate current issues in pathologist shortages in the NHS and worldwide and reduce turnaround times of the screening results. Before deployment in clinical practice, a larger scale validation is required with further analysis of IGUANA’s feature explanation output. In addition, considerable time needs to be invested into extending the current user interface so that it easily integrates with current pathologists’ clinical workflows. This will involve a detailed study on the effectiveness of the decision support tool within abnormal biopsies and assessing its implications on time to report the diagnosis.

## Data Availability

Original WSIs from University Hospitals Coventry and Warwickshire NHS Trust, East Suffolk and North Essex NHS Foundation Trust and South Warwickshire NHS Foundation Trust will be made available upon completion of the PathLAKE project. Relevant information on obtaining the data from the IMP cohort can be found in the original publication. Graph data will be made available upon request, under a non-commercial Creative Commons license, to enable reproduction and improvement of the results obtained in this paper.

## Acknowledgements

We acknowledge Dr SA Sanders and Dr Naresh Chachlani for their assistance in providing WSIs from South Warwickshire NHS Foundation Trust.

## Data Sharing

Original WSIs from University Hospitals Coventry and Warwickshire NHS Trust, East Suffolk and North Essex NHS Foundation Trust and South Warwickshire NHS Foundation Trust will be made available upon completion of the PathLAKE project. Relevant information on obtaining the data from the IMP cohort can be found in the original publication^16^. Graph data will be made available upon request, under a non-commercial Creative Commons license, to enable reproduction and improvement of the results obtained in this paper.

## Contributors

SG, DS and NR designed the study with support from all co-authors. SG, FM and NR developed the methods. SG wrote the code and carried out the experiments. MB provided results using IDaRS and CLAM. MA, YWT, EH, KD, HS, AR, SW, AA, KB, MN, KH, HE, KG and DS provided diagnostic annotations of colonic biopsy slides. SG, FM, MA, KG, DS and NR performed analysis and interpretation of the results. MB, MJ, NW, WL, AB and SEAR provided technical and material support. SG, FM, DS and NR were all involved in the drafting of the paper. NR is the Corresponding Author and guarantor. All authors read and approved the final paper. The corresponding author attests that all listed authors meet authorship criteria and that no others meeting the criteria have been omitted.

## Ethical Approval

This study was conducted under Health Research Authority National Research Ethics approval 15/NW/0843; IRAS 189095 and the Pathology image data Lake for Analytics, Knowledge and Education (PathLAKE) research ethics committee approval (REC reference 19/SC/0363, IRAS project ID 257932, South Central - Oxford C Research Ethics Committee). Data collection and usage of the IMP Diagnostics dataset was performed in accordance with national legal and ethical standards applicable to this cohort^16^.

## Transparency Statement

The lead authors (the manuscript’s guarantors) affirms that the manuscript is an honest, accurate, and transparent account of the study being reported; that no important aspects of the study have been omitted; and that any discrepancies from the study as planned (and, if relevant, registered) have been explained. To help ensure this, the authors followed the principles outlined by the MI-CLAIM^57^ and DECIDE-AI^58^ checklists. We include a completed DECIDE-AI checklist in Supplementary Section S6.

## Supplementary material

### S1. Diagnostic Pathway

**Supplementary Fig 1:**
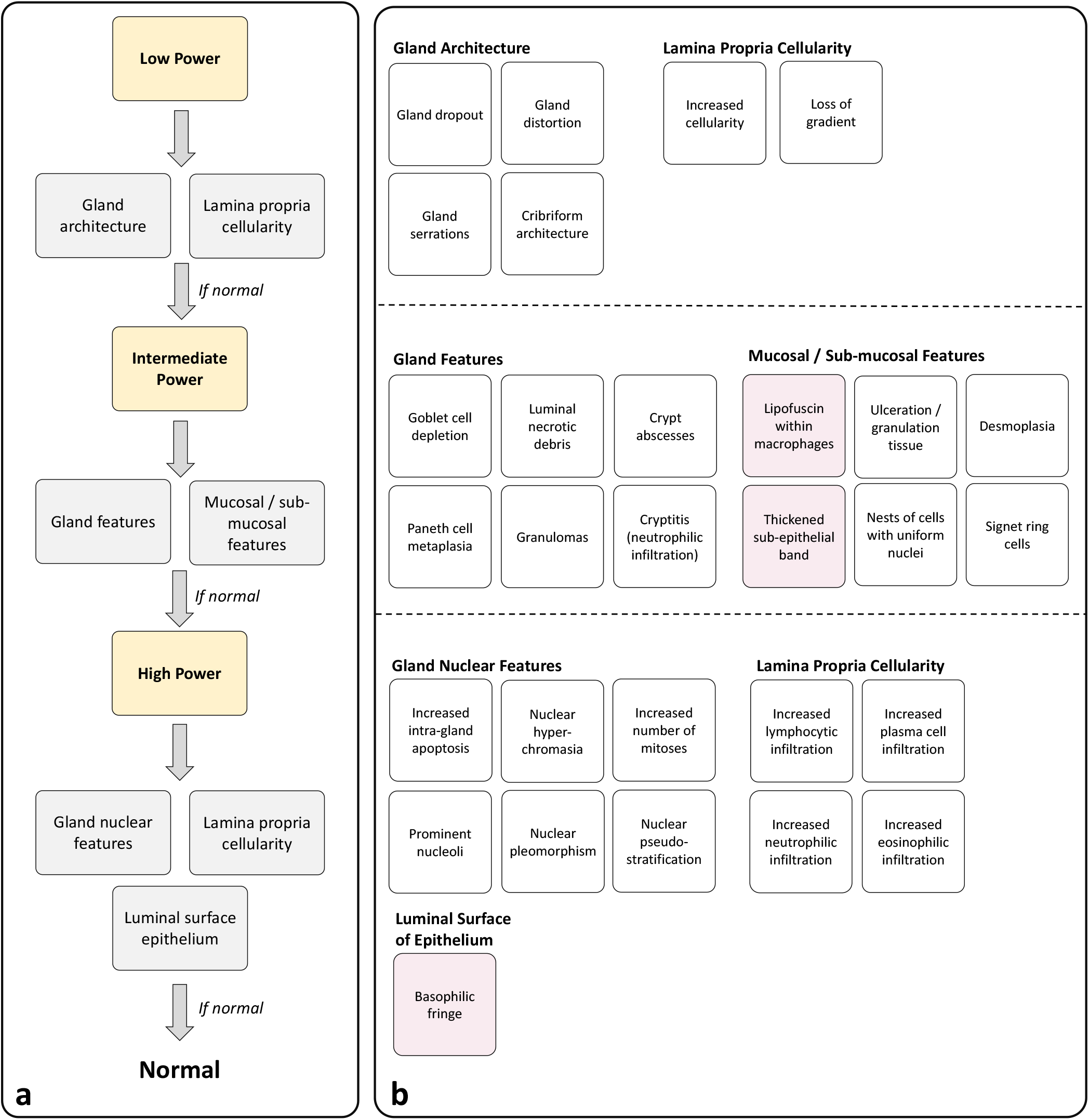
Pathologist colon screening diagnostic algorithm. **a**, Decision process for diagnosing colon biopsies as normal. If any abnormal feature is found during this process, then the entire tissue sample is reported as abnormal. **b**, feature breakdown within each category. Red regions show features not yet explicitly modelled in our approach.

### S2. Data Audit & Cohort Details

**Supplementary Table 1:** Internal audit of seven UK NHS trusts for large bowel biopsies in 2019.

|  | **Histopathology Requests** | **Large Bowel Biopsies (%)** | **Normal Large Bowel Biopsies (%)** |
| --- | --- | --- | --- |
| **Coventry** | 41,771 | 4,877 (11.7) | 1,680 (34.4) |
| **Wolverhampton** | 52,008 | 9,708 (18.7) | 4,140 (42.6) |
| **Oxford** | 56,575 | 7,766 (13.7) | 3,938 (50.7) |
| **Nottingham** | 59,851 | 10,562 (17.6) | 3,428 (32.4) |
| **Newcastle** | 59,843 | 5,348 (8.9) | 2,015 (37.7) |
| **Durham** | 34,958 | 6,240 (17.8) | 2,353 (37.7) |
| **Glasgow** | 108,000 | 29,000 (13.9) | 7,830 (27.0) |
| **Total** | **413,006** | **73,501 (17.8)** | **25,384 (34.5)** |

**Supplementary Table 2:** Summary of all conditions present in the datasets used in the paper.

|  | **UHCW** | **IMP**<br>**Diagnostics** | **ESNE** | **South**<br>**Warwickshire** |
| --- | --- | --- | --- | --- |
| **Normal** | ✓ | ✓ | ✓ | ✓ |
| **Ulcerative Colitis** | ✓ | ✗ | ✓ | ✓ |
| **Collagenous colitis** | ✓ | ✗ | ✓ | ✓ |
| **Crohn's disease** | ✓ | ✗ | ✗ | ✓ |
| **Mild inflammation** | ✓ | ✓ | ✓ | ✓ |
| **Moderate inflammation** | ✓ | ✓ | ✓ | ✓ |
| **Chronic inflammation** | ✓ | ✓ | ✓ | ✓ |
| **CMV** | ✓ | ✗ | ✗ | ✗ |
| **Dysplasia** | ✓ | ✓ | ✓ | ✓ |
| **Hyperplastic polyp** | ✓ | ✓ | ✓ | ✓ |
| **Tubular adenoma** | ✓ | ✓ | ✓ | ✓ |
| **Villous adenoma** | ✓ | ✓ | ✓ | ✓ |
| **Lymphoma** | ✓ | ✗ | ✗ | ✗ |
| **Lipoma** | ✓ | ✗ | ✗ | ✗ |
| **Adenocarcinoma** | ✓ | ✓ | ✓ | ✓ |

**Supplementary Fig 2:**
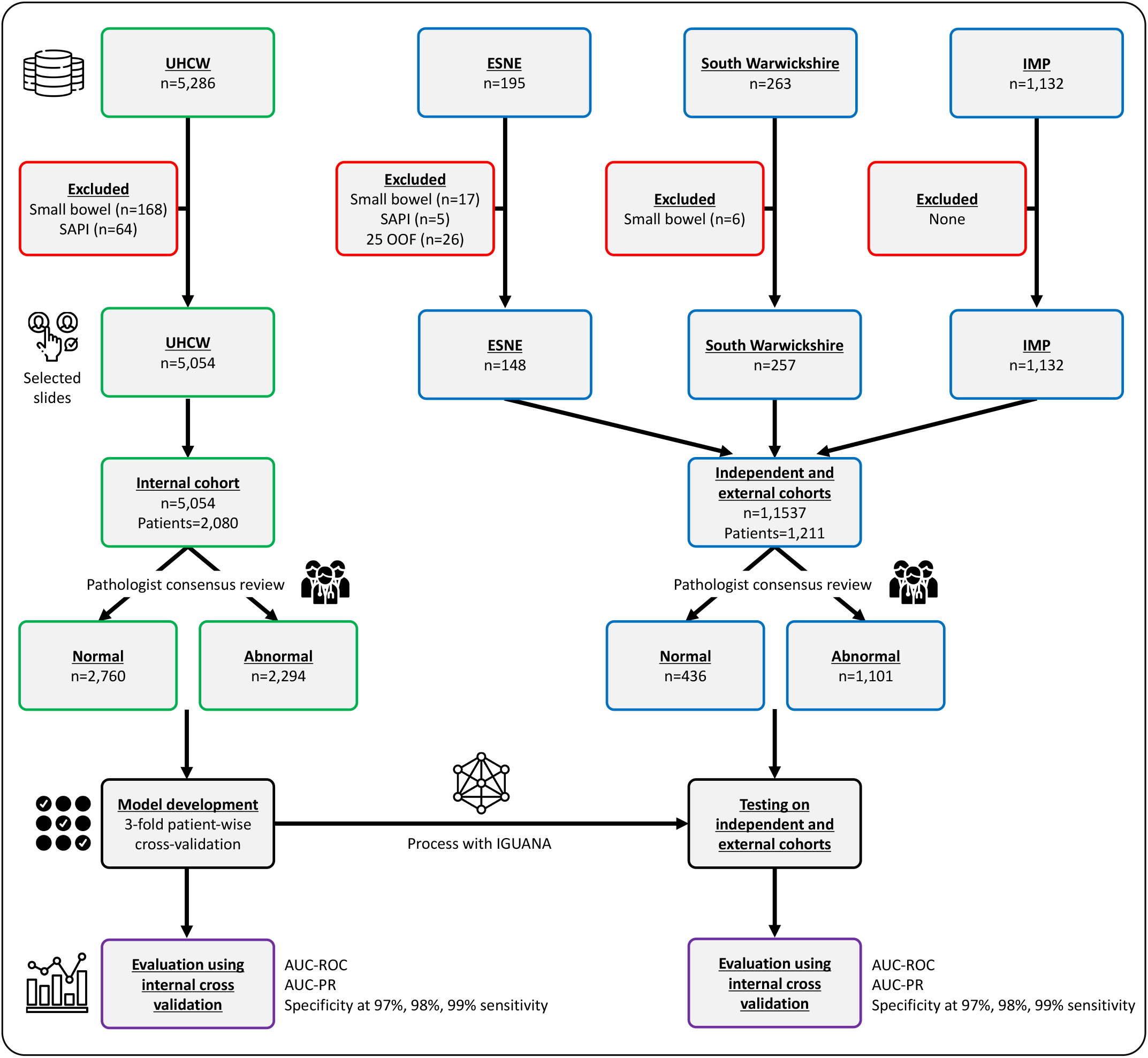
Data description diagram showing the experiment design and the inclusion and exclusion criteria used in the study. Here n denotes the number of whole-slide images, OOF refers to Out-Of-Focus slides and SAPI refers to biopsies that were Slightly Abnormal but Pathologically Insignificant.

**Supplementary Fig 3:**
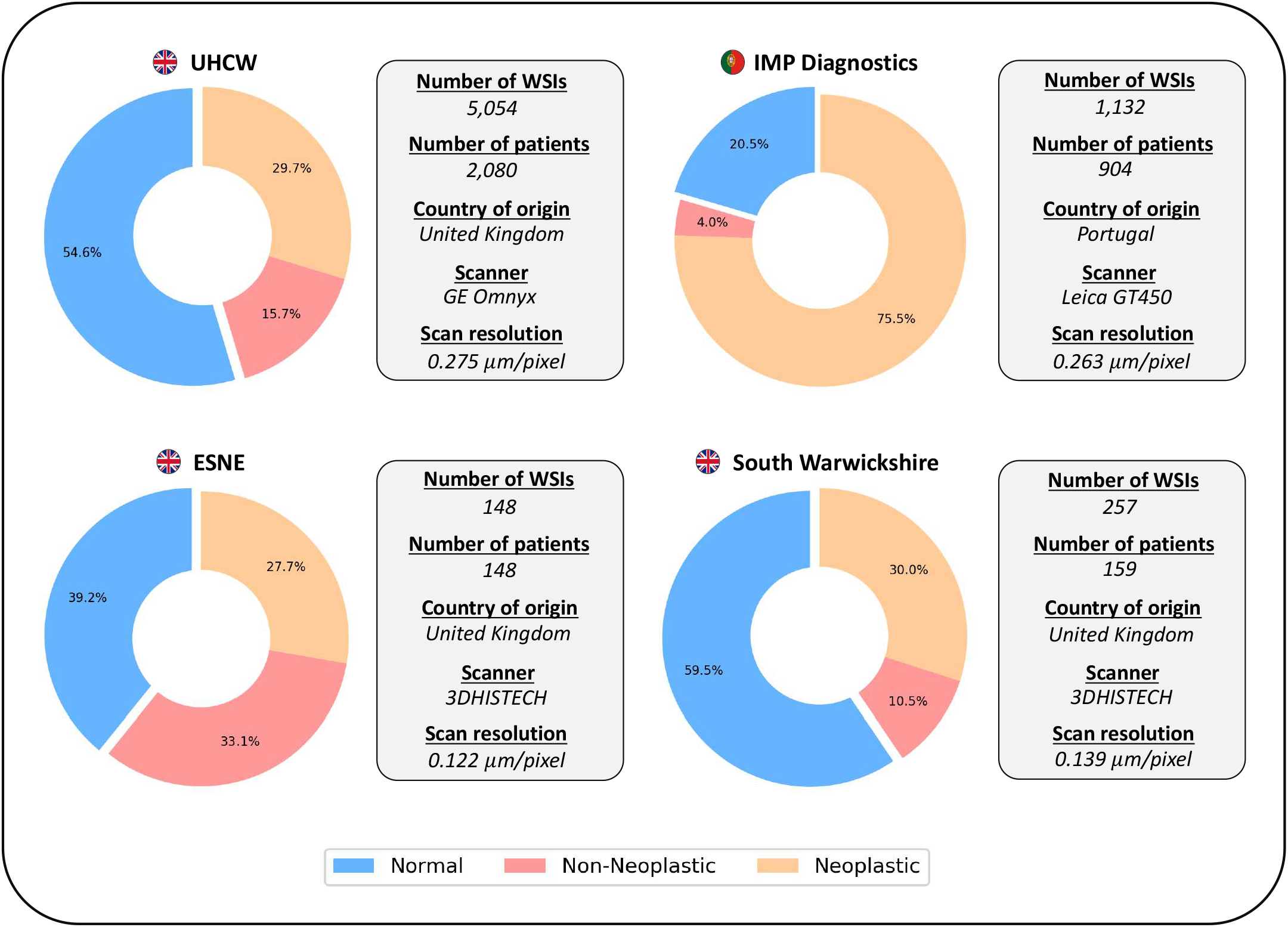
Summary of the data used in this study. Despite using WSIs labelled as either normal or abnormal in our experiments, we also show the breakdown of abnormal slides into non-neoplastic and neoplastic categories.

**Supplementary Fig 4:**
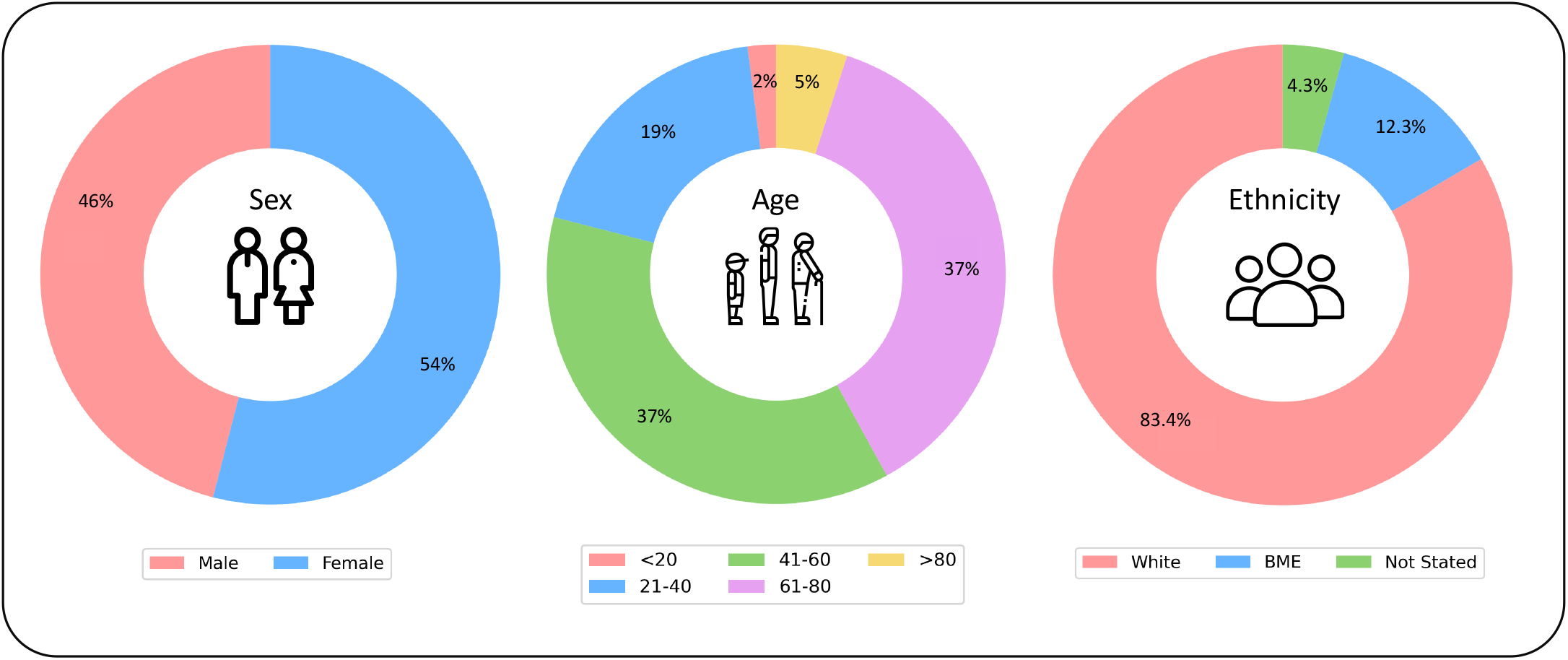
Demographic information of the UHCW dataset, that was used for algorithm development. BME denotes black and minority ethnic groups.

### S3. Extended Methods

#### Graph neural networks for computational pathology

Existing graph neural networks (GNNs) for computational pathology usually consider fixed-size image patches at each node^1-3^ and so fail to incorporate features derived from macrostructures, which can span multiple image patches. However, GNNs that use nodes at centres of image patches in WSIs^4^ may have poor interpretability. Instead, nodes can be centred at known histological entities, such as nuclei and glands, allowing pathologists to directly reason with a model’s predicted outcome^5^. Although some methods position nodes at known entities, Deep Learning-based features are commonly used^6 7^, again leading to reduced interpretability. Rather than using features derived from image regions, graphs built on top of meaningful entities enable the extraction of morphological features. For example, previous methods have constructed graphs on top of nuclei (also known as cell graphs), allowing utilisation of interpretable cellular features^8 9^. However, nuclei are the most basic building blocks in the tissue and therefore associated features may have limited expressive power, and may fail to model important multi-cellular structures, such as glands. Cell graphs can also be very large, where a single tissue sample can contain tens of thousands of nuclei, leading to the generation of intractable graph models. To overcome recent limitations in the literature, our proposed method utilises the concept of gland-graphs for WSI classification, where the nodes are positioned at glands within the tissue, with associated human-interpretable features. The features that we utilise are clinically-meaningful and in line with pathologist diagnostic pathways, leading to excellent performance and providing a highly-interpretable output.

#### Identification of histological objects

Cerberus^10^ is a fully convolutional neural network with a shared ResNet34^11^ encoder and *T* independent decoders, each of which makes a prediction for a specific task *t*. Specifically, we consider 6 tasks: 1) gland instance segmentation, 2) gland semantic segmentation, 3) nuclear instance segmentation, 4) nuclear semantic segmentation, 5) lumen instance segmentation and 6) tissue type patch classification. Here, the instance and semantic segmentation branches are combined to achieve simultaneous segmentation and classification. For segmentation tasks, we use U-Net^12^-inspired decoders that upsample the features step-by-step by a factor of two. After each upsampling operation, we incorporate features from the encoder with skip connections, followed by two convolutions (3×3 kernel) with batch normalisation^13^. This is repeated until the features have the same spatial dimensions as the input. For tissue type classification, we use global average pooling to reduce the features at the output of the encoder to a 256-dimensional vector, which is then followed by two fully connected layers. The tissue type classification output is used to estimate the lamina propria region.

#### Gland-graph construction

Mathematically, a graph is defined as *G* ≡ (*V, E*), where *V* is a set of *N* vertices (or nodes) and *E* is a set of edges, where *e*_*i,j*_ ∈ *E* denotes an edge between nodes *i* and *j* ∈ *V*. In our case, *V* describes the set of all glands in a WSI. Each node typically has an associated *k*-dimensional feature vector ***x***_*i*_ for *i* ∈ *V*. In existing methods, an edge *e*_*i,j*_ is constructed if the Euclidean distance between the centroids of nodes *i* and *j* is less than a certain threshold^9 14^. The distance between neighbouring node centroids is suitable for convex node entities, such as nuclei, because centroids will usually be located within the object. However, glands can often be non-convex, especially when they become cancerous. Therefore, we instead define an edge *e*_*i,j*_ in our gland-graph if the minimum distance between points on the boundary contours of two glands *i* and *j* is less than a certain distance *α*.

#### Gland-graph neural network

Upon formation of our gland-graph representation *G* ≡ (*V, E*) of a WSI in terms of its nodes *V* and their non-directional edges *E*, we pass the input through a GNN to compute the slide-level prediction score. Each node vector ***x***_*i*_ represents a gland in terms of the previously described features and the GNN aggregates information across nodes using the edges in its computation. Note that the number of nodes and edges in each graph can be different depending upon the tissue structure. The GNN first applies a linear operation on ***x***_*i*_ ∈ ℝ^25^ to produce another node-level feature representation 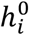 for input into two Principal Neighbourhood Aggregation (PNA) graph convolution^15^ layers. Each PNA layer (*l* = *f*,2) updates each node representation by aggregating information from its neighbours *j* ∈ *ℵ*_*i*_ according to the following rule: 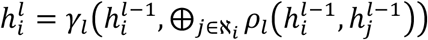, where *γ*_*l*_ and *ρ*_*l*_ are multi-layer perceptrons (MLPs) each with their own trainable weights. PNA uses a combination of aggregation strategies (denoted by ⊕) based on scaling of mean, standard deviation, minimum and maximum aggregation operators over node features. It has been shown recently that using this aggregation approach is superior to methods that use a single aggregation step, such as computing the sum as it enables the resulting GNN to be a better discriminator of local graph structures^15^. Outputs of the two PNA layers are concatenated and fused with a linear operation to arrive at the final node-level feature embedding ***r***_*i*_. The final output ***f***(*G*) ∈ ℝ^*C*^ is obtained by performing global attention pooling:

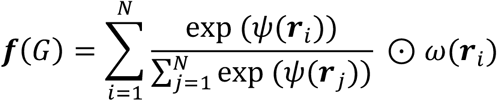

where *ψ* and *ω* are MLPs, *C* denotes the number of classes predicted by the network, ⊙ denotes element-wise multiplication and hence the global pooling operator learns to assign a varying weight to different gland representations, signifying their relative importance in the final prediction. Finally, a softmax function is applied and all the trainable weights in the GNN are optimised in an end-to-end fashion by minimising the binary cross entropy loss between the output and the ground-truth labels of training slides.

#### Gland-graph explanation

To enable an interpretable and explainable output, we utilise the graph pruning method GNNExplainer^16^, which generates a subset of nodes and features that play a crucial role in the GNN’s prediction. The intuition here is that removing unimportant nodes and features should have a negligible impact on the performance and can therefore be removed. Specifically, GNNExplainer is formulated as an optimisation task that maximises the mutual information between a GNN’s prediction and the distribution of possible subgraph structures. Practically, this is achieved by learning a real-valued mask, which gives less weight to unimportant graph components. For our approach, we learn a node explanation mask *M*_*n*_ ∈ ℝ^*N*^ and a feature explanation mask *M*_*f*_ ∈ ℝ^*N*×25^, where *N* denotes the number of nodes in each WSI and 25 is the pre-defined number of features. Rather than applying a threshold to the learned masks to give a compact subgraph, we visualise the raw mask output, which gives an interpretable and explainable output that can be discussed with clinicians. Specifically, the learned mask *M*_*n*_ provides the node explanation which can be overlaid on top of the glands in each WSI as a heatmap. Similarly, *M*_*f*_ can be used to identify the top features for each node/gland and the corresponding importance values. To obtain a WSI-level explanation, the local features are averaged within the top ten most predictive nodes.

To assess which node explanation method was best, we utilised the metric proposed by Jaume *et al*.^*5*^. The intuition behind their proposed metric is that a superior node explanation technique should be able to locate top nodes that can better differentiate between classes (in our case normal *vs* abnormal). We compared node explanations given by GNNExplainer, integrated gradients^31^ and the attention scores given by our network and found that GNNExplainer gave the best results in terms of class separability. It is important to note that GNNExplainer is a *post-hoc* method, which is why we used attention pooling during optimisation of our predictive model.

#### Comparative methods

To rigorously assess the performance of our approach, we compare with IDaRS^17^, CLAM^18^ and a random forest classifier using the interpretable glandular features that we extracted (denoted by Gland-RF). Both IDaRS and CLAM are recent state-of-the-art DL approaches that use raw H&E image patches as input in a multiple-instance learning (MIL) framework. The Gland-RF model computes the mean and standard deviation of all local features within the slide to obtain a fixed-size global feature vector before input to the model. When fitting the Gland-RF for each fold, we perform a grid search over the hyperparameters and select the best models in terms of their performance on the validation set.

### S4. Interpretable Features

**Supplementary Fig 5:**
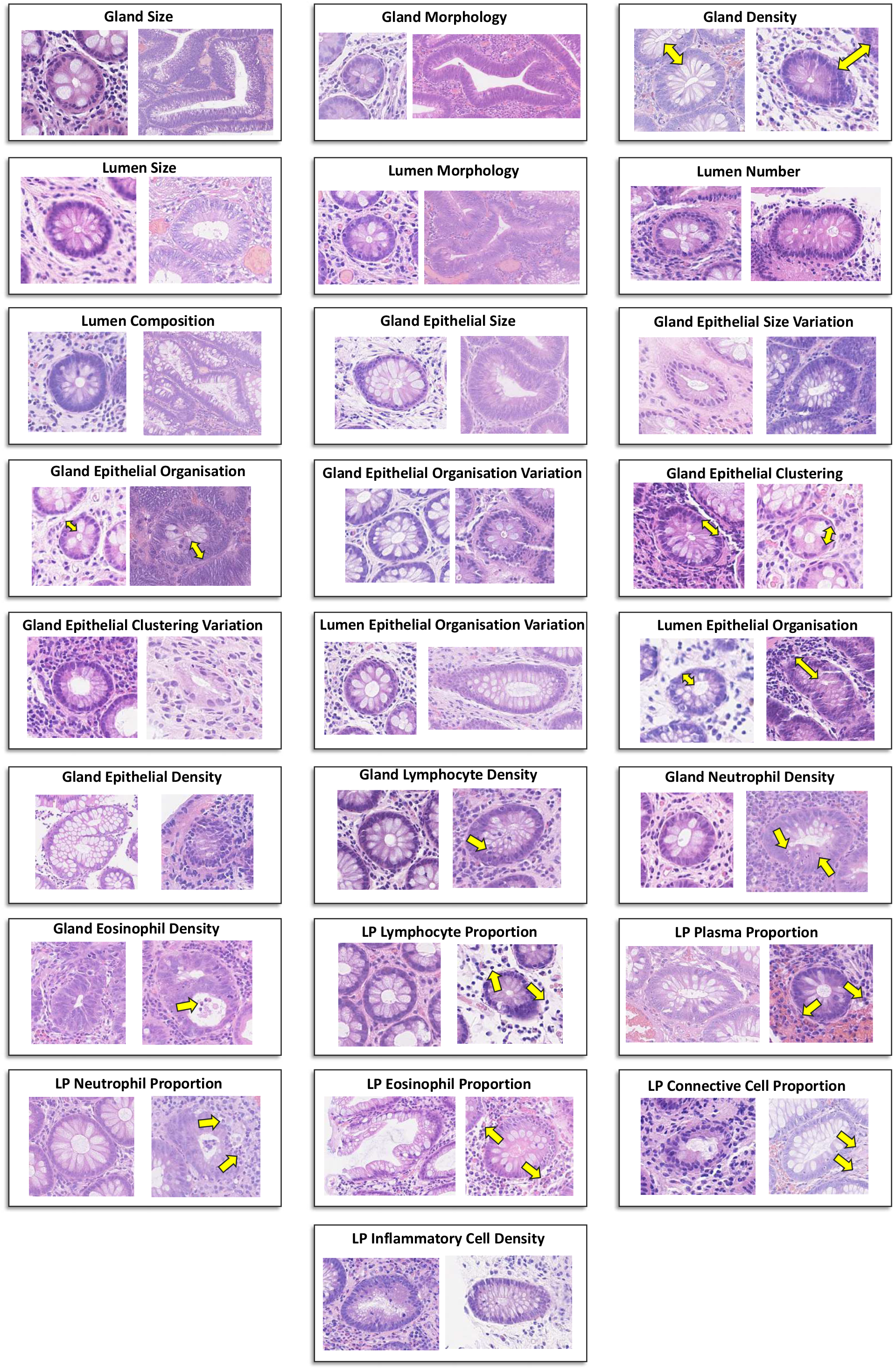
Examples of cropped image regions from the UHCW dataset containing features taken from the 5^th^ (first image in each panel) and 95^th^ (second image in each panel) percentiles. Yellow arrows show areas within the image relevant to the associated feature.

**Supplementary Table 3:** Description of all 25 features used in our experiments, along with various conditions that they may be able to detect. LP denotes lamina propria.

| Feature Name | Feature Description | Histological Description | Main Conditions Modelled |
| --- | --- | --- | --- |
| Gland size | Size of gland (number of pixels at 0.5 microns/pixel) | Gland enlargement | Neoplasia, dysplasia, adenomatous polyps |
| Gland morphology | How far gland is from being elliptical – BAM distance<sup>38</sup> | Gland distortion, gland branching | Neoplasia, dysplasia, adenomatous polyps |
| Gland density | Distance to nearest gland | Gland dropout | Inflammation |
| Lumen size | Size of lumen (number of pixels at 0.5 microns/pixel) | Lumen dilation | Neoplasia, dysplasia, hyperplastic polyps |
| Lumen morphology | How far lumen is from being elliptical – BAM distance<sup>38</sup> | Lumen serrations | Hyperplastic polyps |
| Lumen number | Lumen count within gland | Cribriform architecture | Neoplasia |
| Lumen composition | Ratio of lumen to gland area | Gland dilation | Hyperplasia |
| Gland epithelial size | Average size of epithelial nuclei within a gland | Epithelial cell atypia | Neoplasia, dysplasia, adenomatous polyps |
| Gland epithelial size variation | Standard deviation of epithelial nuclei size within a gland | Epithelial cell atypia | Neoplasia, dysplasia, adenomatous polyps |
| Gland epithelial organisation | Average distance of intra-gland epithelial nuclei to nearest gland boundary | Stratification of epithelial cells | Neoplasia, dysplasia, adenomatous polyps |
| Gland epithelial organisation variation | Standard deviation of intra-gland epithelial nuclei distances to nearest gland boundary | Uneven stratification of epithelial cells | Neoplasia, dysplasia, adenomatous polyps |
| Gland epithelial clustering | Average distance between intra-gland epithelial nuclei | Epithelial cells tightly packed | Neoplasia, dysplasia, adenomatous polyps |
| Gland epithelial clustering variation | Standard deviation of intra-gland epithelial nuclei distances to nearest gland boundary | Epithelial cells unevenly spaced | Neoplasia, dysplasia, adenomatous polyps |
| Lumen epithelial organisation | Average distance of intra-gland epithelial nuclei to nearest lumen boundary | Gland dilation, cribriform architecture | Neoplasia, dysplasia, hyperplastic polyps |
| Lumen epithelial organisation variation | Standard deviation of intra-gland epithelial nuclei distances to nearest lumen boundary | Lumen serrations, cribriform architecture | Neoplasia, dysplasia, hyperplastic polyps |
| Gland epithelial density | Number of intra-gland epithelial nuclei, normalised by the gland size | Solid sheets of epithelial cells | Neoplasia, dysplasia, adenomatous polyps |
| Gland lymphocyte density | Number of intra-gland lymphocytes, normalised by the gland size | Gland lymphocyte infiltration | Inflammation |
| Gland neutrophil density | Number of intra-gland neutrophils, normalised by the gland size | Gland neutrophil infiltration (crypt abscess) | Inflammation |
| Gland eosinophil density | Number of intra-gland eosinophils, normalised by the gland size | Gland eosinophil infiltration | Inflammation |
| LP lymphocyte proportion | Proportion of lymphocytes within nearest 220 nuclei to gland | Lymphocytic colitis | Inflammation |
| LP plasma cell proportion | Proportion of plasma cells within nearest 220 nuclei to gland | Colitis | Inflammation |
| LP neutrophil proportion | Proportion of neutrophils within nearest 220 nuclei to gland | Acute inflammation | Inflammation |
| LP eosinophil proportion | Proportion of eosinophils within nearest 220 nuclei to gland | Eosinophilic colitis | Inflammation |
| LP connective tissue cell proportion | Proportion of connective tissue cells within nearest 220 nuclei to gland | Desmoplasia | Inflammation |
| LP inflammatory cell density | Mean distance of nearest 250 inflammatory nuclei to gland | General inflammation | Inflammation, hyperplastic polyps |

### S5. Extended Results

**Supplementary Fig 6:**
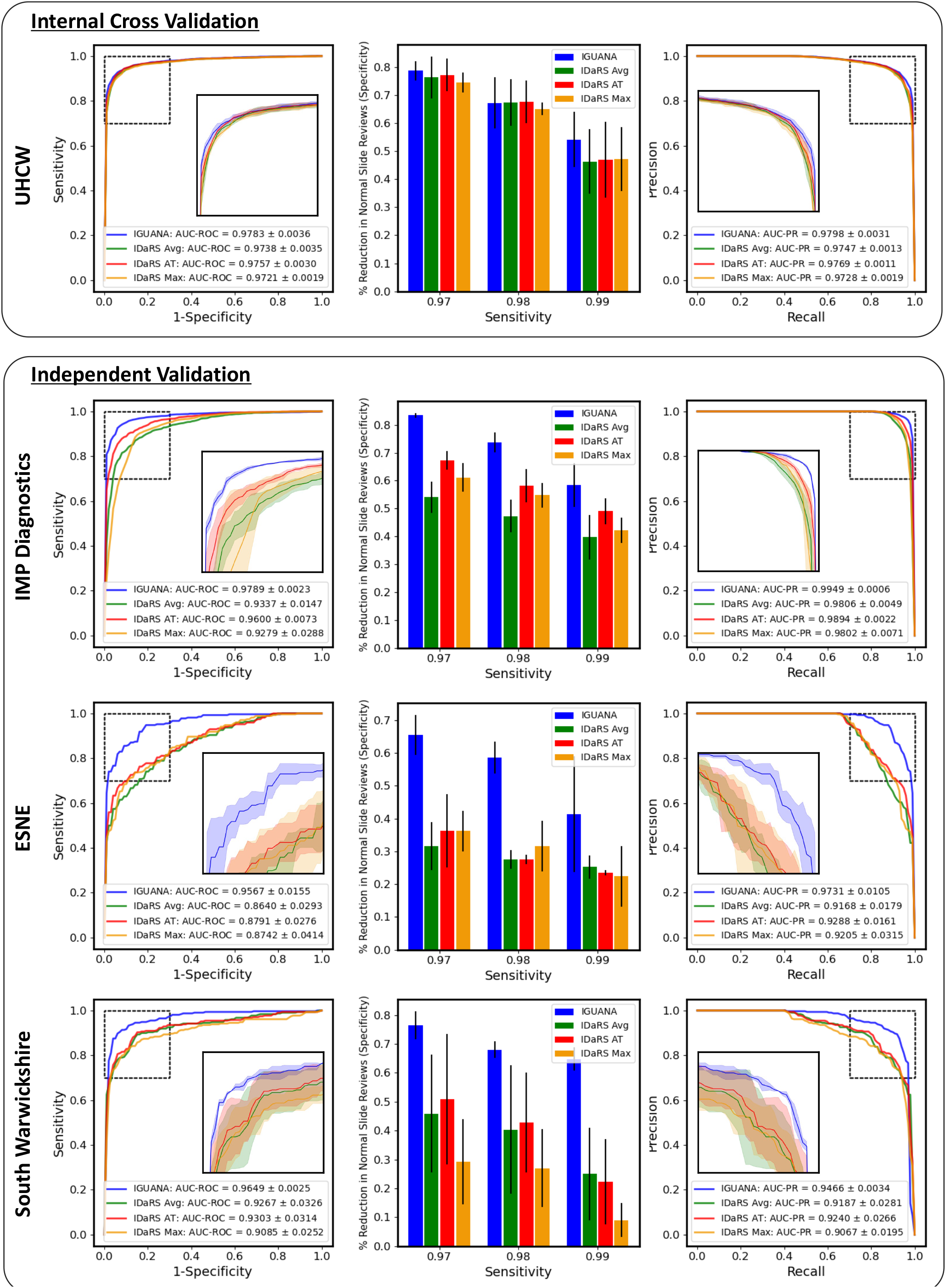
Results of IGUANA across the four cohorts used in our experiments compared to IDaRS with different aggregation strategies. Here, AT corresponds to the average of top tiles, where top tiles are those with a score above the median. We display the ROC and PR curves along with the respective AUC scores for each method. We also display the specificities obtained at sensitivity cut-offs of 0.97, 0.98 and 0.99. The shaded areas in the curves and the error bars in the bar plots show 1 standard deviation from the results.

**Supplementary Fig 7:**
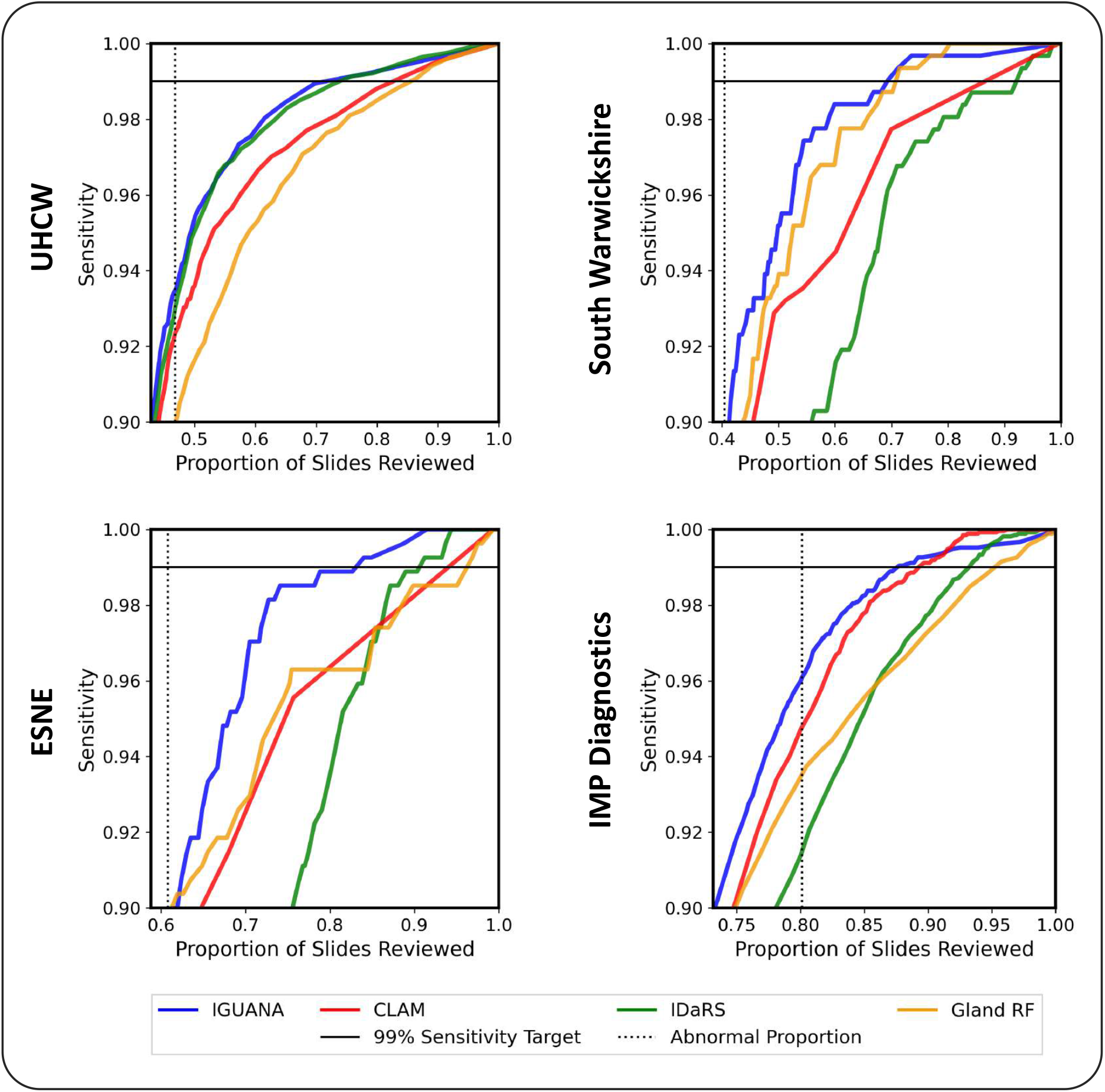
Impact of the automatic colon biopsy screening tool on clinical practice. For each dataset we show the proportion of slides that need to be reviewed to ensure a specific sensitivity. Our target sensitivity is 0.99. We also show the proportion of abnormal slides to indicate the minimum number of slides that need to be reviewed to ensure high sensitivity.

**Supplementary Fig 8:**
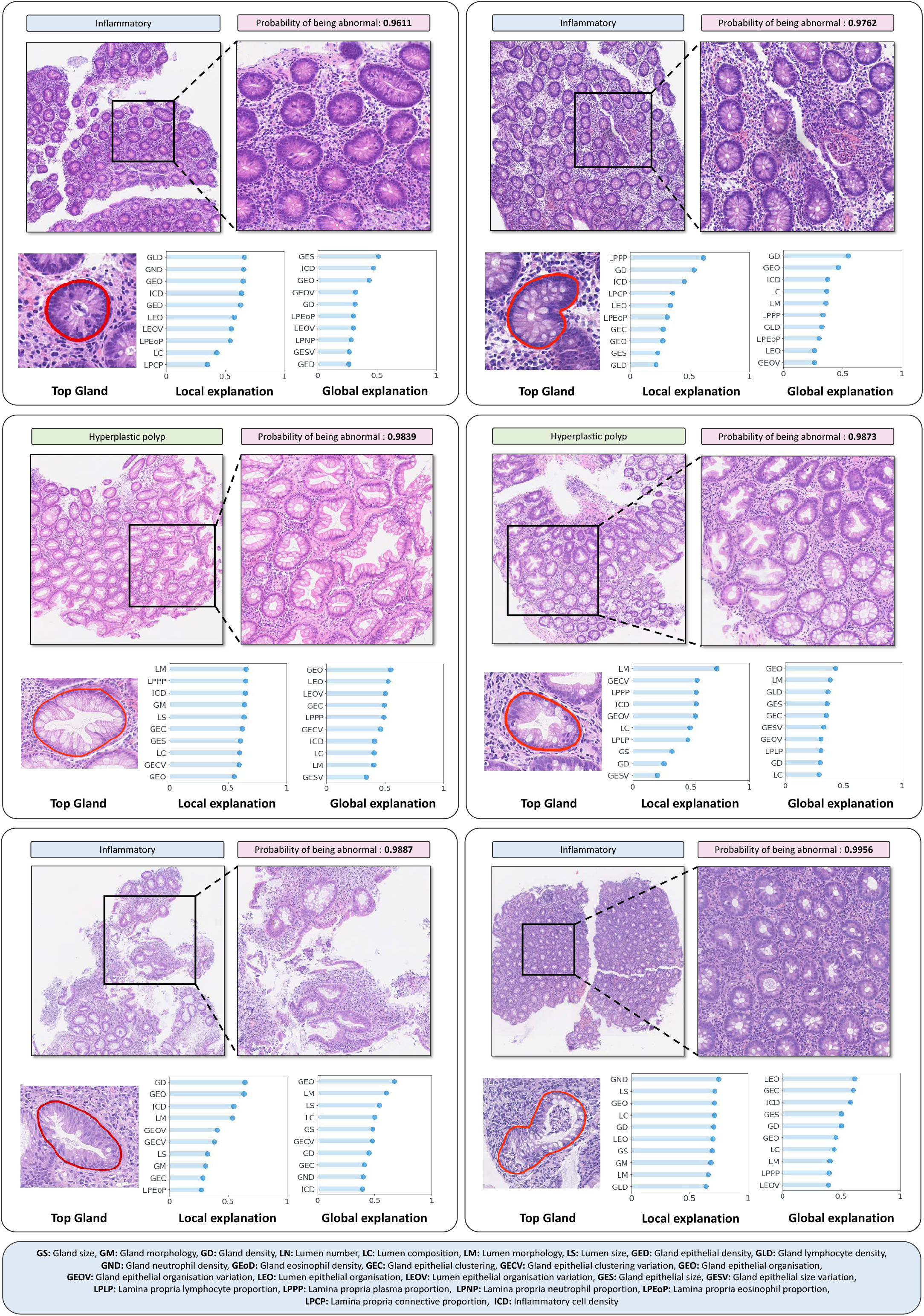
Analysis of false positives in the UHCW dataset. In each panel, we display zoomed in images, the gland that contributes most to the prediction, the local feature explanation (corresponding to the top gland) and the global feature explanation.

**Supplementary Fig 9:**
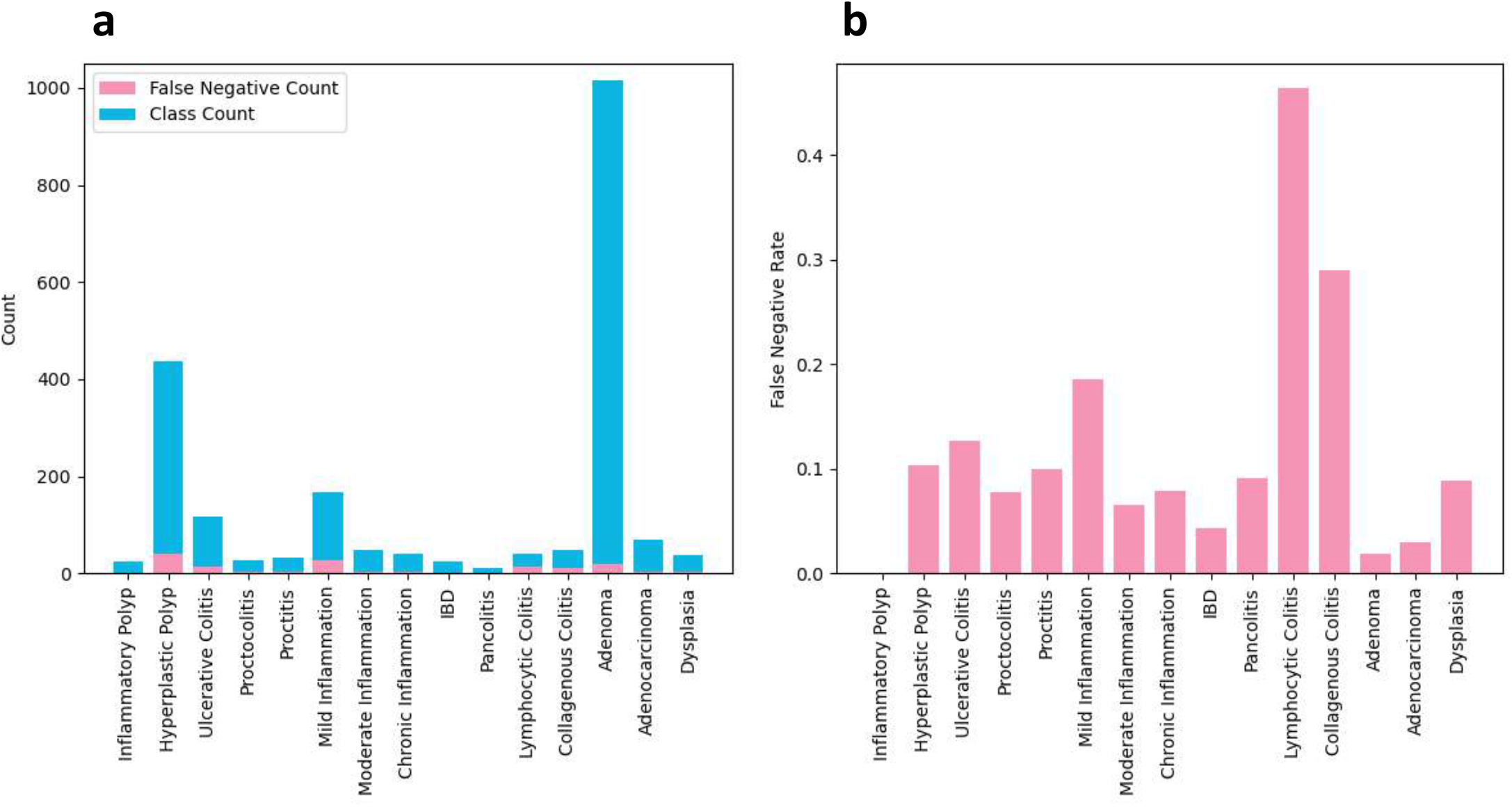
False negative analysis. On the left we show the class counts along with the corresponding number of false negatives. On the right, we show the false positive rate per class. For this figure, we don’t consider sub-conditions with minimal examples.

### S6. DECIDE-AI Checklist

**Supplementary Table 4:**
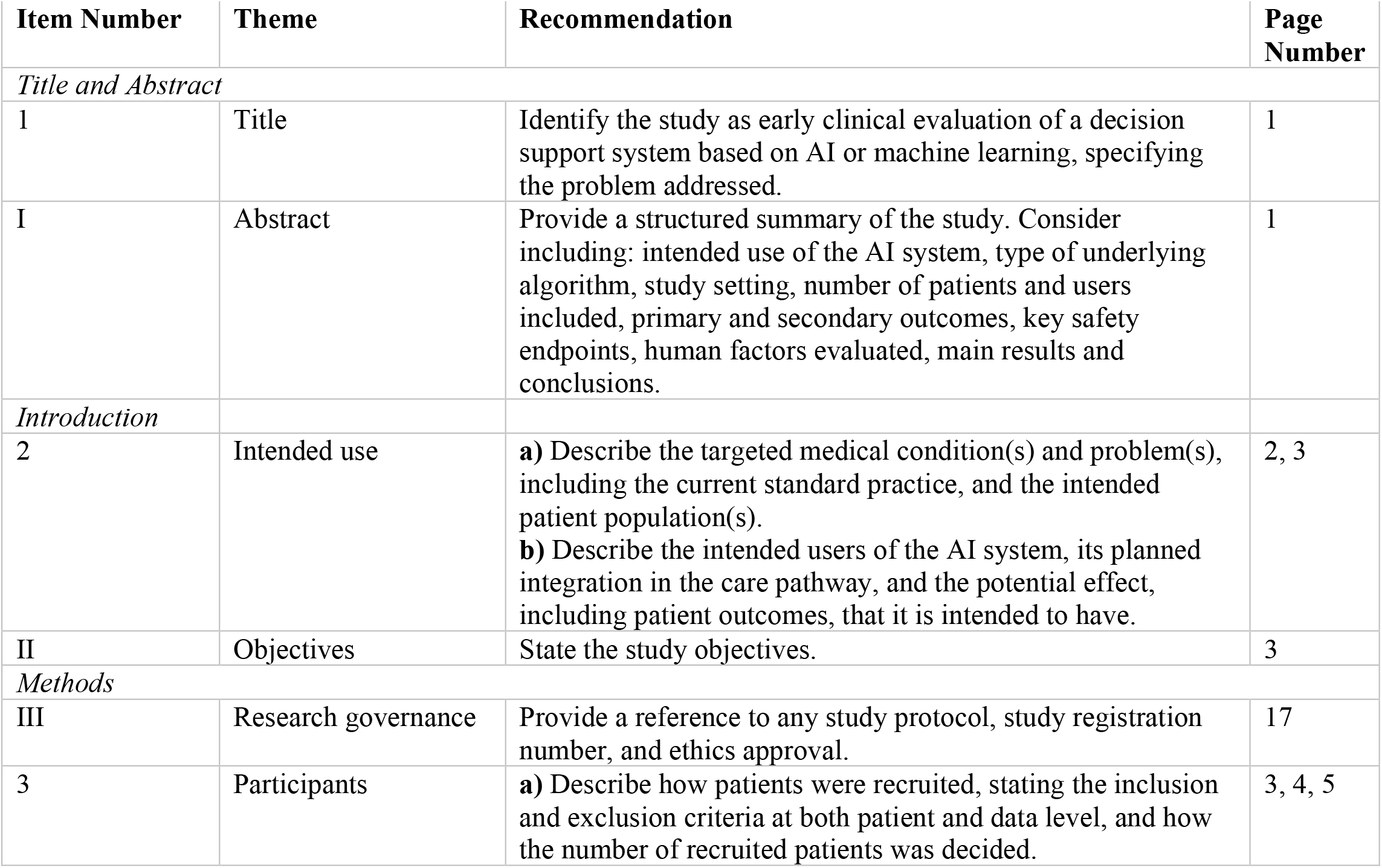
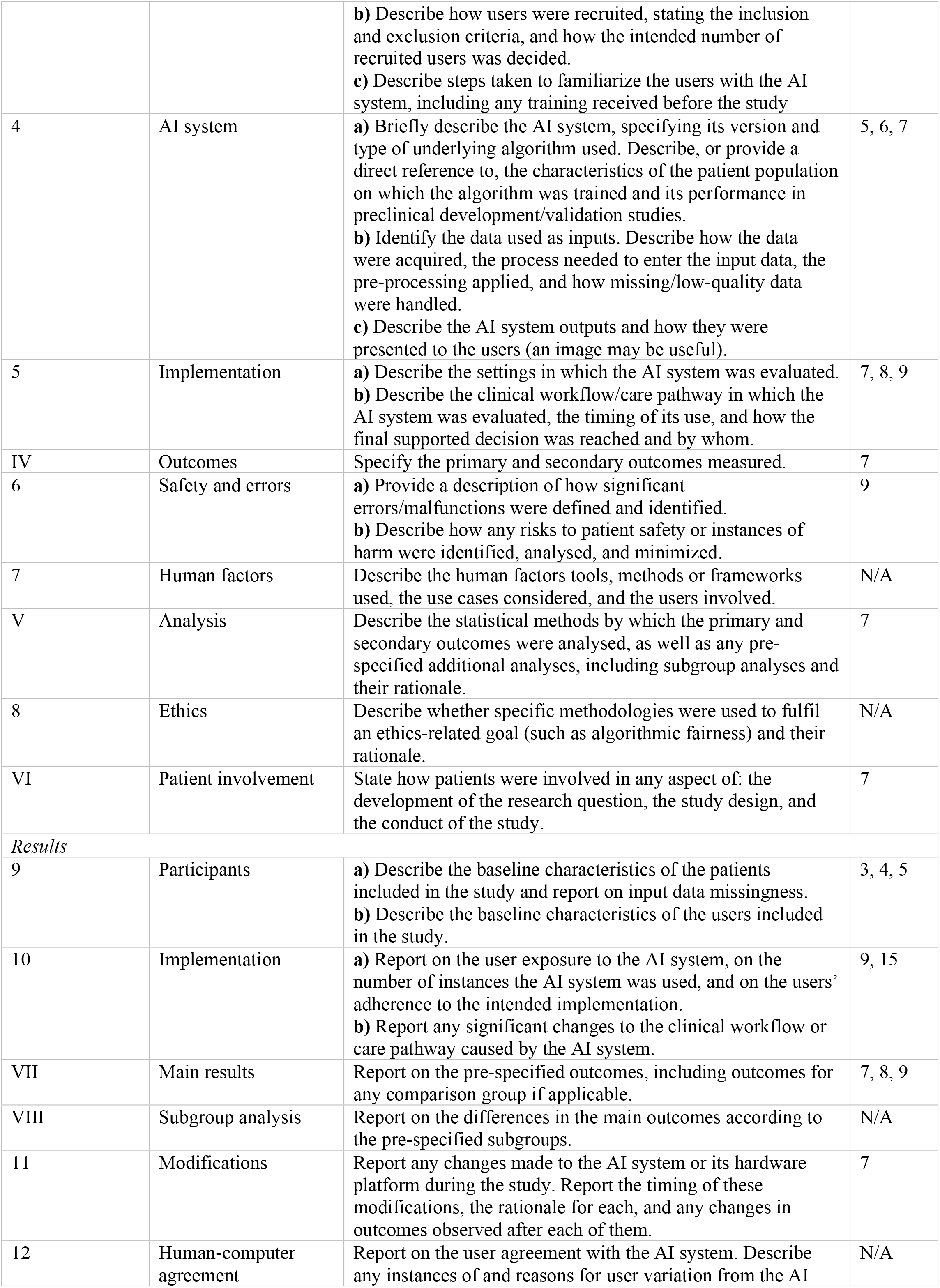
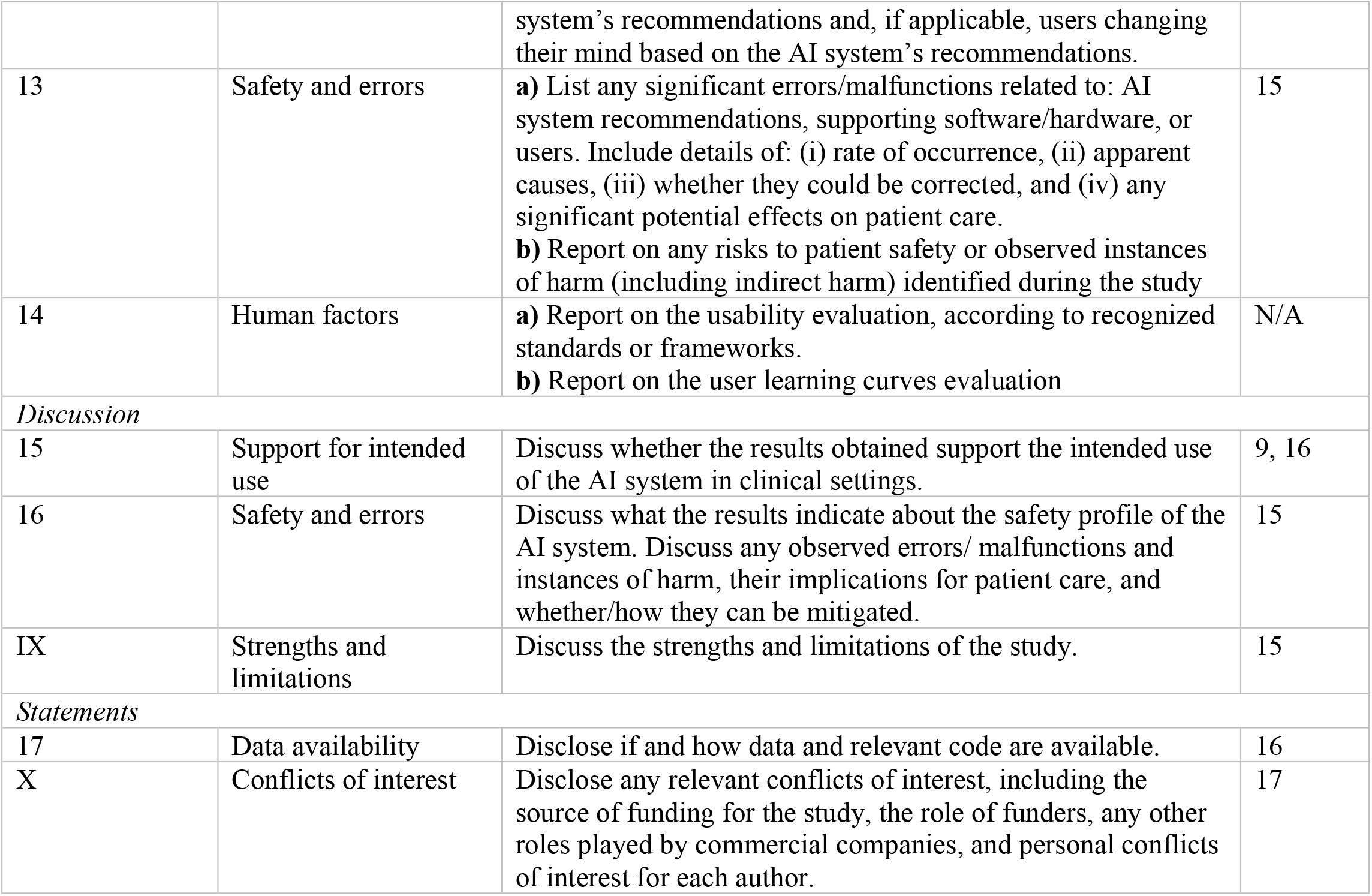
DECIDE-AI checklist^19^ indicating where each item was adhered to in the main manuscript. In this study, there was no human interaction with the AI tool and hence those items are not applicable. 1-14 denote AI-specific reporting items and I-X denote genetic reporting items.

| Item Number | Theme | Recommendation | Page Number |
| --- | --- | --- | --- |
| *Title and Abstract* |  |  |  |
| 1 | Title | Identify the study as early clinical evaluation of a decision support system based on AI or machine learning, specifying the problem addressed. | 1 |
| I | Abstract | Provide a structured summary of the study. Consider including: intended use of the AI system, type of underlying algorithm, study setting, number of patients and users included, primary and secondary outcomes, key safety endpoints, human factors evaluated, main results and conclusions. | 1 |
| *Introduction* |  |  |  |
| 2 | Intended use | **a)** Describe the targeted medical condition(s) and problem(s), including the current standard practice, and the intended patient population(s).<br>**b)** Describe the intended users of the AI system, its planned integration in the care pathway, and the potential effect, including patient outcomes, that it is intended to have. | 2, 3 |
| II | Objectives | State the study objectives. | 3 |
| *Methods* |  |  |  |
| III | Research governance | Provide a reference to any study protocol, study registration number, and ethics approval. | 17 |
| 3 | Participants | **a)** Describe how patients were recruited, stating the inclusion and exclusion criteria at both patient and data level, and how the number of recruited patients was decided. | 3, 4, 5 |
|  |  | **b)** Describe how users were recruited, stating the inclusion and exclusion criteria, and how the intended number of recruited users was decided.<br>**c)** Describe steps taken to familiarize the users with the AI system, including any training received before the study |  |
| 4 | AI system | **a)** Briefly describe the AI system, specifying its version and type of underlying algorithm used. Describe, or provide a direct reference to, the characteristics of the patient population on which the algorithm was trained and its performance in preclinical development/validation studies.<br>**b)** Identify the data used as inputs. Describe how the data were acquired, the process needed to enter the input data, the pre-processing applied, and how missing/low-quality data were handled.<br>**c)** Describe the AI system outputs and how they were presented to the users (an image may be useful). | 5, 6, 7 |
| 5 | Implementation | **a)** Describe the settings in which the AI system was evaluated.<br>**b)** Describe the clinical workflow/care pathway in which the AI system was evaluated, the timing of its use, and how the final supported decision was reached and by whom. | 7, 8, 9 |
| IV | Outcomes | Specify the primary and secondary outcomes measured. | 7 |
| 6 | Safety and errors | **a)** Provide a description of how significant errors/malfunctions were defined and identified.<br>**b)** Describe how any risks to patient safety or instances of harm were identified, analysed, and minimized. | 9 |
| 7 | Human factors | Describe the human factors tools, methods or frameworks used, the use cases considered, and the users involved. | N/A |
| V | Analysis | Describe the statistical methods by which the primary and secondary outcomes were analysed, as well as any pre-specified additional analyses, including subgroup analyses and their rationale. | 7 |
| 8 | Ethics | Describe whether specific methodologies were used to fulfil an ethics-related goal (such as algorithmic fairness) and their rationale. | N/A |
| VI | Patient involvement | State how patients were involved in any aspect of: the development of the research question, the study design, and the conduct of the study. | 7 |
| *Results* |  |  |  |
| 9 | Participants | **a)** Describe the baseline characteristics of the patients included in the study and report on input data missingness.<br>**b)** Describe the baseline characteristics of the users included in the study. | 3, 4, 5 |
| 10 | Implementation | **a)** Report on the user exposure to the AI system, on the number of instances the AI system was used, and on the users' adherence to the intended implementation.<br>**b)** Report any significant changes to the clinical workflow or care pathway caused by the AI system. | 9, 15 |
| VII | Main results | Report on the pre-specified outcomes, including outcomes for any comparison group if applicable. | 7, 8, 9 |
| VIII | Subgroup analysis | Report on the differences in the main outcomes according to the pre-specified subgroups. | N/A |
| 11 | Modifications | Report any changes made to the AI system or its hardware platform during the study. Report the timing of these modifications, the rationale for each, and any changes in outcomes observed after each of them. | 7 |
| 12 | Human-computer agreement | Report on the user agreement with the AI system. Describe any instances of and reasons for user variation from the AI system's recommendations and, if applicable, users changing their mind based on the AI system's recommendations. | N/A |
| 13 | Safety and errors | **a)** List any significant errors/malfunctions related to: AI system recommendations, supporting software/hardware, or users. Include details of: (i) rate of occurrence, (ii) apparent causes, (iii) whether they could be corrected, and (iv) any significant potential effects on patient care.<br>**b)** Report on any risks to patient safety or observed instances of harm (including indirect harm) identified during the study | 15 |
| 14 | Human factors | **a)** Report on the usability evaluation, according to recognized standards or frameworks.<br>**b)** Report on the user learning curves evaluation | N/A |
| *Discussion* |  |  |  |
| 15 | Support for intended use | Discuss whether the results obtained support the intended use of the AI system in clinical settings. | 9, 16 |
| 16 | Safety and errors | Discuss what the results indicate about the safety profile of the AI system. Discuss any observed errors/ malfunctions and instances of harm, their implications for patient care, and whether/how they can be mitigated. | 15 |
| IX | Strengths and limitations | Discuss the strengths and limitations of the study. | 15 |
| *Statements* |  |  |  |
| 17 | Data availability | Disclose if and how data and relevant code are available. | 16 |
| X | Conflicts of interest | Disclose any relevant conflicts of interest, including the source of funding for the study, the role of funders, any other roles played by commercial companies, and personal conflicts of interest for each author. | 17 |

